# Immune and behavioral correlates of protection against symptomatic post-vaccination SARS-CoV-2 infection

**DOI:** 10.1101/2023.08.25.23294626

**Authors:** Emilie Goguet, Cara H. Olsen, William A. Meyer, Sara Ansari, John H. Powers, Tonia L. Conner, Si’Ana A. Coggins, Wei Wang, Richard Wang, Luca Illinik, Margaret Sanchez Edwards, Belinda M. Jackson-Thompson, Monique Hollis-Perry, Gregory Wang, Yolanda Alcorta, Mimi A. Wong, David Saunders, Roshila Mohammed, Bolatito Balogun, Priscilla Kobi, Lakeesha Kosh, Kimberly Bishop-Lilly, Regina Z. Cer, Catherine E. Arnold, Logan J. Voegtly, Maren Fitzpatrick, Andrea E. Luquette, Francisco Malagon, Orlando Ortega, Edward Parmelee, Julian Davies, Alyssa R. Lindrose, Hannah Haines-Hull, Matthew S. Moser, Emily C. Samuels, Marana S. Tso, Elizabeth K. Graydon, Allison M.W. Malloy, David R. Tribble, Timothy H. Burgess, Wesley Campbell, Sara Robinson, Christopher C. Broder, Robert J. O’Connell, Carol D. Weiss, Simon Pollett, Eric D. Laing, Edward Mitre

## Abstract

We sought to determine pre-infection correlates of protection against SARS-CoV-2 post-vaccine infections (PVI) acquired during the first Omicron wave in the United States. Serum and saliva samples from 176 vaccinated adults were collected from October to December of 2021, immediately before the Omicron wave, and assessed for SARS-CoV-2 Spike-specific IgG and IgA binding antibodies (bAb). Sera were also assessed for bAb using commercial assays, and for neutralization activity against several SARS-CoV-2 variants. PVI duration and severity, as well as risk and precautionary behaviors, were assessed by questionnaires. Serum anti-Spike IgG levels assessed by research assay, neutralization titers against Omicron subvariants, and low home risk scores correlated with protection against PVIs after multivariable regression analysis. Commercial assays did not perform as well as research assay, likely due to their lower dynamic range. In the 32 participants that developed PVI, anti-Spike IgG bAbs correlated with lower disease severity and shorter duration of illness.

## 1. Introduction

Since the emergence of severe acute respiratory syndrome coronavirus 2 (SARS-CoV-2), the global pandemic of coronavirus 19 disease (COVID-19) has caused substantial morbidity and mortality ^1^. Identifying reliable correlates of protection with quantitative antibody concentrations against clinically relevant outcomes, such as symptomatic SARS-CoV-2 infection, has been a major scientific goal throughout the pandemic. Robust correlates would enable policy makers to better determine boosting strategies, facilitate assessment of novel SARS-CoV-2 vaccines (including against emerging viral variants), and could potentially be used to guide patient-specific interventions such as repeat boosting, prophylactic antibody administration, or enhanced non-pharmaceutical interventions in immunocompromised or other high-risk individuals ^2–4^.

To date, both post-vaccination binding antibodies (bAb) and neutralizing titers (NT) have been associated with protection against SARS-CoV-2 infection in several studies ^2,4–17^, but the strength of correlation and the levels associated with protection vary substantially between such reports ^18^. Challenges involved in identifying a reliable and reproducible correlate include heterogeneous populations, differences in infectiousness of SARS-CoV-2 variants over time, continually changing community-wide transmission levels, different individual exposure risks, and variability and evolution of individual risk mitigation behaviors. To our knowledge, prior studies have not been able to incorporate all of these factors into adjusted correlate of protection analyses. Finally, the exact outcome used for defining protection (e.g., any infection, symptomatic infection, severe infection) can also differ.

In this study, we sought to identify correlates of protection against symptomatic post-vaccine infections (PVI) in a cohort of vaccinated, generally healthy adults during the first Omicron wave in the United States. To enhance our ability to identify such correlates, we utilized serum and saliva samples that were obtained from participants just before the initial Omicron wave in the U.S., evaluated individual risk exposures and mitigation behaviors, and tested Wuhan-1 wild-type (WT), D614G and Omicron-specific bAb levels and NTs. Additionally, we made comparisons between our research-based bAb assay (MMIA) with two commercially available assays for anti-receptor binding domain (RBD) bAb, and we used a validated symptom questionnaire to assess if potential correlates of protection are associated with decreased severity or duration of symptoms in individuals that experienced symptomatic PVI. We demonstrate an adaptable analytical framework that incorporates patient demography and risk behavior into immune correlate of protection estimates.

## 2. Methods

### 2.1. Study participants

Details of the PASS study protocol have been previously published ^19^. Inclusion criteria included being ≥ 18 years of age, generally healthy, and working at the Walter Reed National Military Medical Center (WRNMMC). Exclusion criteria included being immunocompromised, history of COVID-19 infection, and seropositivity for WT anti-Spike (S) IgG bAb at time of screening. The PASS study was initiated in August 2020, with rolling enrollment through March 2021. The study protocol was approved by the Uniformed Services University Institutional Review Board and all participants provided informed consent. Monthly research clinic visits to obtain serum and saliva were scheduled until August 2021, after which the visits were scheduled quarterly until August 2022. The subset of participants included for analysis here had received at least 2 doses of mRNA vaccine by December 1, 2021 and were seen at the research clinic between October 1 and December 15, 2021 (Fall 2021 visit). The observation period ran from after the Fall 2021 research clinic visit to April 1, 2022, with the first PVI occurring on December 1, 2021.

### 2.2. Collection of viral respiratory infection symptoms

At the Spring 2022 research clinic visit, study participants that had experienced a PVI completed a validated viral respiratory infection patient-reported outcome symptom questionnaire (FLU-PRO Plus ©), as described before ^20,21^. Briefly, FLU-PRO Plus measures severity, frequency and duration of 34 symptoms organized in 7 symptom domains: nasal, throat, body/systemic, chest, sense (taste/smell), gastrointestinal, and eyes. The severity of each is measured on a scale from 0 (absent) to 4 (greatest intensity), then mean scores in each symptom domain are summed for a total FLU-PRO Plus symptom score (0 to 28). Each participant is also asked about the duration of symptoms experienced (reported in days).

### 2.3. Diagnosis of SARS-CoV-2 infection

Participants were asked to obtain PCR and/or antigen testing whenever they experienced any symptoms consistent with an upper respiratory infection. Results and methods of testing were reported to the research clinic during the Spring 2022 visit.

### 2.4. Risk exposure and precautionary measures behavioral assessments

Risk exposure and adherence to precautionary measures, in the workplace and at home, was also assessed for each participant. A behavioral questionnaire for the Winter 2021 period was completed by participants at the Spring 2022 research clinic visit. The questionnaire is divided into 4 categories: work exposure risks, work precautionary measures, home exposure risks, and home precautionary measures. Each category has between 3 to 5 questions, with most questions being worth between 0 (did not experience that exposure or did not use that precautionary measure) to 4 points (did experience that risk exposure or used that precautionary measure all the time). Scores for each category were reported on a 0 – 100 scale **(Supplementary Table 1)**.

### 2.5. Research anti-S binding antibody testing

Blood samples were collected in SST tubes (BD Biosciences, Franklin Lakes, NJ, US) during the Fall 2021 research clinic visit and aliquots of serum were frozen at −80°C until day of testing. Samples were tested for IgG and IgA bAbs against SARS-CoV-2 Spike (S) protein of the Wuhan-1 WT strain and the BA.1 Omicron subvariant, using a microsphere-based multiplex immunoassay (MMIA) built using Luminex xMAP-based technology. The S protein of the WT strain and the BA.1 Omicron subvariant, expressed as native-like prefusion stabilized ectodomain trimers, were sourced from Curia (Albany, NY, US) and Dr. Dominic Esposito (Protein Expression Laboratory, NCI FNL, US), respectively. Trimeric S protein antigens (Ag) were coupled to magnetic carboxylated microspheres (Luminex, Austin, TX, US) as previously described ^22^. On day of testing, serum samples were thawed at room temperature and then thermally-inactivated for 30 minutes at 60°C. Individual serum samples were tested at 1:400, 1:8000, 1:16,000, and 1:32,000 dilutions in PBS and mixed with S protein-coupled microspheres. Biotinylated cross-adsorbed goat anti-human IgG (Invitrogen cat# A18815) was used for detection of anti-S bound IgG and biotinylated cross-adsorbed goat anti-human IgA (Invitrogen cat# A18791) was used for detection of anti-S bound IgA. Antigen-antibody complexes were incubated with streptavidin-phycoerythrin (BioRad, Hercules, CA, US) and measured as a median fluorescence intensity (MFI) by a Bio-Plex 200 HTF multiplex system (BioRad, Hercules, CA, US). IgG and IgA bAb concentrations were interpolated against an internal standard curve of pooled serum from nine individuals with previous SARS-CoV-2 infection. IgG bAb levels for anti-S (WT) were reported in WHO Binding Antibody Units/ml (BAU/ml) after calibration of the internal standard to the U.S. Human SARS-CoV-2 Serology Standard ^23^. Anti-S (BA.1) IgG and anti-S (WT) IgA were reported in arbitrary binding units (AU/ml).

### 2.6. Commercial binding antibody testing

Serum samples collected during the Fall 2021 research clinic visit were tested using the US Food and Drug Administration Emergency Use Authorization (FDA EUA)-cleared Elecsys® anti-SARS-CoV-2 S assay (Roche Diagnostics, Indianapolis, IN, US) and the ADVIA Centaur® SARS-CoV-2 IgG (sCOVG) assay (Siemens Healthcare Diagnostics Inc., Tarrytown, NY, US). Both assays are semi-quantitative.

The Elecsys® anti-SARS-CoV-2 S assay (for use on the cobas e analyzers) uses a recombinant protein representing the RBD of the S antigen (Ag) in a double-sandwich assay format. The Ag within the reagent capture predominantly anti-SARS-CoV-2 IgG, but also capture anti-SARS-CoV-2 IgA and IgM. Thus, the assay measures total bAb levels (IgG, IgA, and IgM) against the RBD of WT S protein. Results are expressed in Units (U)/ml. An additional 1:10 dilution (resulting in total 1:200 dilution) was performed on all serum samples, as most of the initial 1:20 dilutions tested were above the upper limit of the analytical measuring interval (0.40-250 U/ml). Roche-specific U were converted to WHO BAU by the following formula: BAU = Roche U/ 0.972.

The ADVIA Centaur® sCOVG assay is an automated chemiluminescent sandwich immunoassay that measures IgG bAb levels against WT SARS-CoV-2 RBD in serum. The system reports the assay results in Index Values (0.5 to 100). The SARS-CoV-2 IgG bAb levels were then reported in WHO BAU/ml using the following formula: BAU/ml = 45.078 x Index Value(0.7984).

### 2.7. Saliva binding antibody testing

Saliva samples were collected using the passive drool method (saliva collection aid from Salimetrics, Centre County, PA, US) frozen at −80°C immediately following collection. On day of testing, samples were thawed on ice then centrifuged at 16,000 x g for 10 minutes, at 4°C. The supernatant was then transferred to a new Eppendorf tube and the remaining pellet stored at - 80°C for future testing. The saliva supernatants were heat-inactivated for 30 minutes at 60°C and then diluted 1:5 in PBS before proceeding with the modified MMIA initially developed for serum samples. Saliva samples were tested for IgG, IgA, and secretory IgA (sIgA) bAbs against the WT SARS-CoV-2 S protein. Detection antibodies for IgG and IgA were the same as in the serum MMIA, and a cross-adsorbed goat anti-human sIgA (MyBioSource cat# MBS534541) biotinylated using a biotinylation kit (Abcam cat# ab201795) was used for detection of sIgA. Arbitrary binding units (AU/ml) for salivary bAb were calculated by interpolating MFI values against standard curves generated using microspheres conjugated with known concentrations of human IgG, IgA, or sIgA.

### 2.8. Pseudovirus production and neutralization assay

HIV-based lentiviral pseudoviruses with desired SARS-CoV-2 S proteins (D614G, B.1.617.2, BA.1 and BA.1.1) were generated as previously described ^24^. Pseudovirus neutralization assays were performed using 293T-ACE2-TMPRSS2 cells in 96-well plates ^25^. Pseudoviruses with titers of approximately 106 relative luminescence units per milliliter (RLU/mL) of luciferase activity were incubated with serially diluted sera for 2 hours at 37°C prior to inoculation onto the plates that were pre-seeded one day earlier with 3.0 x 10^4^ cells/well. Pseudovirus infectivity was determined 48 hours post-inoculation for luciferase activity by luciferase assay reagent (Promega, Madison, WI, US) according to the manufacturer’s instructions. The inverse of the sera dilutions causing a 50% reduction of RLU compared to control was reported as the neutralization titer (ID_50_). Titers were calculated using a nonlinear regression curve fit (GraphPad Prism Software Inc., La Jolla, CA, US). The mean titer from at least 2 independent experiments each with intra-assay duplicates was reported as the final titer.

### 2.9. Sequencing and bioinformatics

RNA was extracted from the available nasopharyngeal swabs performed at WRNMMC for COVID-19 diagnosis. SARS-CoV-2-specific amplicon sequencing libraries were prepared using the NEBNext ARTIC SARS-CoV-2 Library Prep Kit (New England Biolabs, Ipswich, MA, US) as recommended by the manufacturer. Quality control of the resulting sequencing libraries, consisting of DNA concentration and fragment sizes, was conducted using the Qubit dsDNA HS Assay kit (Thermo Fisher Scientific, Waltham, MA, US) and Agilent High Sensitivity DNA kit (Agilent, Santa Clara, CA, US), respectively. Multiplexed sequencing was performed on the Illumina MiSeq platform (Illumina, San Diego, CA, US) using 600 cycle v3 chemistry. Viral Amplicon Illumina Workflow 2.3 (VAIW) was used to collate and analyze SARS-CoV-2 genomes from the resulting sequencing reads and generate final consensus genomes when possible ^26^. Single Nucleotide Variants (SNVs) were determined using SAMtools mpileup and iVar (intrahost variant analysis of replicates) ^27^. Thresholds were frequency of 0.3 or above, minimum alternate allele read depth of 10 or above, and Q20 or above Phred score to reduce calling false positives. Lineage determination of consensus genomes was conducted using Pangolin (Phylogenetic Assignment of Named Global Outbreak LINeages; v4.1.2) ^28^. Nextstrain clades were determined by Nextclade CLI 2.4.0, Nextalign CLI 1.10.1. Nextstrain overall sequence QC scores of ‘bad’, ‘mediocre’, and ‘good’ were translated into ‘low’, ‘medium’, and ‘high’ for confidence of clade assignment.

### 2.10. Statistical analysis

Means and standard deviations (SD) were calculated using GraphPad Prism 9.4.1 software. Mann-Whitney test was used for unpaired comparisons, with the Bonferroni correction applied for analyses with multiple comparisons. For each assay, antibody levels were plotted against the incidence of infection in individuals above that level. Multivariable logistic regression analysis was performed to determine the effect of the different immunological factors on the risk of post-vaccination infections. Odds ratios relating each assay to infection status were estimated first without adjustment and after adjusting for age, sex, and home risk score. Age and sex were selected *a priori* as covariates and home risk score was included as it was identified as the strongest behavioral predictor of infection. Continuous independent variables were rescaled so that the odds ratio corresponds to a change of 1/10 of the range of the variable. Predictive accuracy of regression models was assessed using the area under the ROC curve and goodness-of-fit was assessed using the Hosmer-Lemeshow test. The Spearman rank correlation was used to described the association between symptom variables and antibody levels. Regression models and McFadden PseudoR2 values were estimated using STATA version 15.

### 2.11. Data availability

Data for this study are available from the Infectious Disease Clinical Research Program (IDCRP), headquartered at the Uniformed Services University of the Health Sciences (USU), Department of Preventive Medicine and Biostatistics. Review by the USU Institutional Review Board is required for use of the data collected under this protocol. Furthermore, the data set includes Military Health System data collected under a Data Use Agreement that requires accounting for uses of the data. Data requests may be sent to: Address: 6270A Rockledge Drive, Suite 250, Bethesda, MD 20817.

## 3. Results

### 3.1. Study Participants’ Demographics

A total of 271 participants were initially enrolled in the PASS study between August 25, 2020 and March 31, 2021. For this analysis, we initially selected all 203 PASS participants that attended the scheduled Fall 2021 research clinic visit (between October 1, 2021 and December 15, 2021) that occurred just before the first Omicron wave in the United States. Of these, 27 participants were excluded from analysis because they received a COVID-19 booster dose during the study’s observation period (between the Fall 2021 research clinic visit and April 1, 2022) **(Supplementary Figure 1)** and thus the Fall 2021 research clinic visit antibody levels would not reflect the most recent titers they had during the Omicron wave.

Of these 176 participants, 123 (69.9%) were female and 53 (30.1%) were male, with a median age of 42.5 (range: 20 – 70; interquartile range [IQR]: 34.0 – 52.0) years. The distribution of race was 71.0% White, 13.1% Black, 9.1% Asian, 4.5% others, and 2.3% not reported. Thirty-two (18.2%) of the 176 participants were diagnosed with a SARS-CoV-2 post-vaccination infection (PVI), whether by PCR or self-collected antigen testing. Of these, 17 (53.1%) were female and 15 (46.9%) were male, with a median age of 44.5 (range: 20 – 63; IQR: 32.8 – 50.3) years, and the racial composition was 62.5% White, 21.9% Black, 6.2% Asian, and 9.4% others. The uninfected group was comprised of 144 participants, 106 (73.6%) of whom were female and 38 (26.4%) were male. The median age was 42 (range: 21 – 70; IQR: 34.0 – 52.0) and the racial composition was 72.9% White, 11.1% Black, 9.7% Asian, 3.5% others, and 2.8% not reported. The majority of the study cohort had a low Charlson Comorbidity Index (CCI) of 0 or 1 (87.5% and 7.4%, respectively when considering the whole cohort). The repartition of CCIs within the uninfected group was fairly similar to the whole cohort (85.4% of 0, 9% of 1, 4.2% of 2, and 1.4% of 3, while the PVI group had 31 participants with an index of 0 (96.9%) and 1 participant with an index of 2 (3.1%) **(Table 1)**.

**Table 1.**
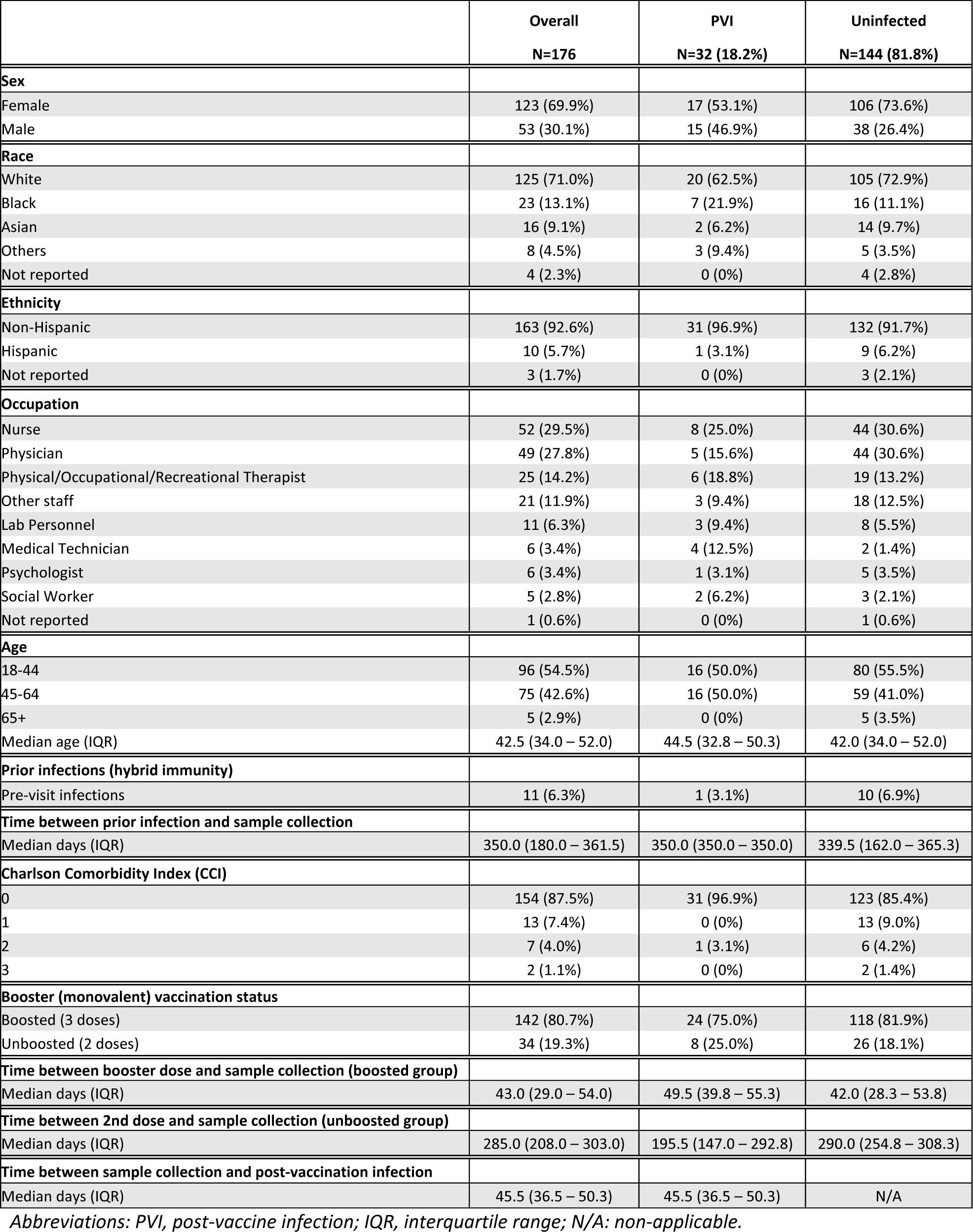
Characteristics of the study cohort.

Study participants represented a wide array of apparently healthy hospital employees with nurses (29.5%), physicians (27.8%), and physical/occupational/recreational therapists (14.2%) making up the majority of the cohort. The same general distribution can be seen for the 32 PVI participants (25.0% nurses, 15.6% physicians, and 18.8% physical/occupational/recreational therapists) **(Table 1)**.

### 3.2. COVID-19 vaccination and SARS-CoV-2 post-vaccine infection history among the study cohort

All of the 176 participants had received at least 2 doses of ancestral monovalent BioNTech/Pfizer BNT162b2 mRNA vaccine for their initial immunization regimen. Of the 176 participants, 142 (80.7%) were boosted (3 ancestral monovalent mRNA vaccine doses) and 34 (19.3%) had only received 2 doses of ancestral monovalent mRNA vaccine. Of the 142 boosted participants, 140 (98.6%) participants received the ancestral monovalent BioNTech/Pfizer BNT162b2 mRNA vaccine booster, and 2 (1.4%) received the ancestral monovalent Moderna mRNA-1273 vaccine booster; both of these participants were in the uninfected group (data not shown). Eleven individuals (6.3%) had hybrid immunity at time of the Fall 2021 visit, with one (3.1%) in the PVI group and 10 (6.9%) in the uninfected group **(Table 1)**.

Thirty-two participants were diagnosed with SARS-CoV-2 PVI during the observation period that started after the Fall 2021 research clinic visit (October to December 2021) and finished on April 1, 2022. Twenty-six (81.3%) were diagnosed by PCR testing and 6 (18.7%) were diagnosed by self-collected antigen testing (data not shown).

Eluents of nasopharyngeal swabs were available for sequencing from 13 of the 32 PVI participants, and all of the samples produced coding complete genomes. Omicron subvariantBA.1.1 was identified in 6 (46.1%) participants, Omicron subvariants BA.1.18 and BA.1.20 were each identified in 2 (15.4%) participants, and Omicron subvariants BA.1.15, BA.1.19, and Delta variant AY.25 were each identified in 1 (7.7%) participant **(Supplementary Table 2)**. Inferring variants by date of infection using GSAID data for the participants in whom sequencing was unavailable ^29^, we estimate that in total 29 of the 32 (90.6%) PVIs were with Omicron variants and 3 (9.4%) were with Delta variants.

Of the 32 participants in the PVI group, 24 (75.0%) were mRNA vaccine-boosted and 8 (25.0%) were not boosted. The distribution of boosted individuals was higher in the uninfected group (n=144), with 118 (81.9%) boosted participants and 26 (18.1%) unboosted participants **(Table 1)**.

### 3.3. Pre-Omicron wave binding IgG serum levels correlate with protection against post-vaccine infections

Serum samples were obtained during the Fall 2021 research clinic visit, at a median of 52.5 (IQR: 45.8 – 67.3) days after last immunization for the PVI group (participants, N=32; serum samples available for analysis, n=31), and 46.5 (IQR: 32.8 – 58.3) days after last immunization for the uninfected group (participants, N=144; serum samples available for analysis, n=140).

The geometric mean (GM) of anti-S (WT) IgG bAb levels measured by MMIA was significantly greater in those who remained uninfected compared to those who developed PVI (3,863 vs 2,736 BAU/ml, p=0.0098) **(Figure 1A)**. When looking at the distribution of anti-S (WT) IgG bAb serum levels, the percentage of PVIs among the study participants decrease from 23.3% and 29.6% for levels of <2,500 BAU/ml and 2,500-5,000 BAU/ml, respectively, to 7.4% at 5,000-7,500 BAU/ml, 8.3% at 7,500-10,000 BAU/ml, and 9.5% at 10,000-15,000 BAU/ml. No infections were detected in participants with levels >15,000 BAU/ml **(Figure 1B, Supplementary Figure 2A)**. Lower serum levels of anti-S (WT) IgA bAb in the PVI group also were observed with a GM of 9,134 AU/ml versus 11,940 AU/ml in the uninfected group, but this difference was not statistically significant (p=0.1167) **(Figure 1C)**. No PVIs were observed at serum anti-S (WT) IgA levels greater than 80,000 AU/ml **(Figure 1C, Supplementary Figure 2D**).

**Figure 1.**
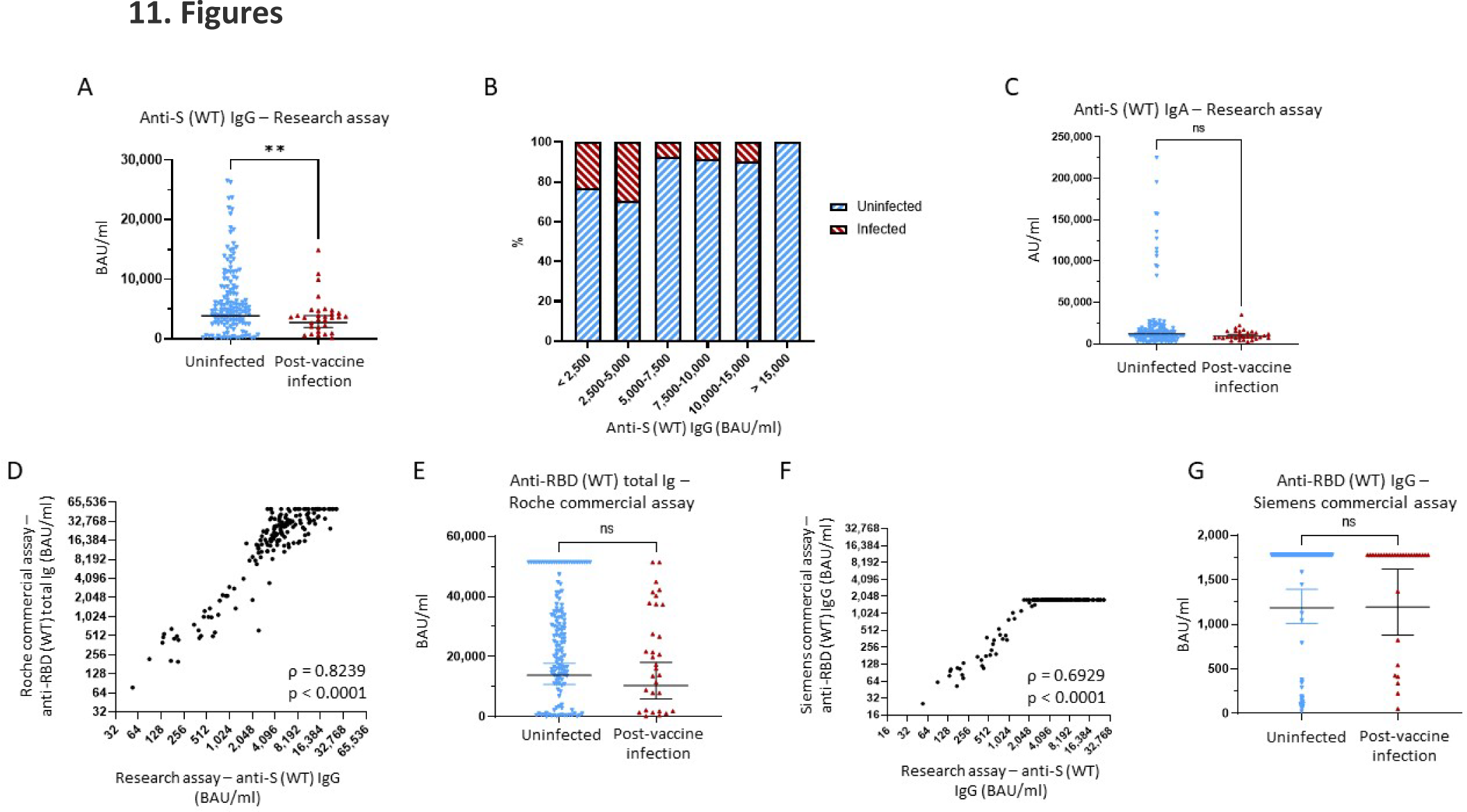
Binding antibody serum levels against Wuhan-1 wild-type (WT) Spike in research and commercial assay. **(A)** Comparison of anti-Wuhan-1 wild-type (WT) Spike (S) IgG serum levels between the uninfected group and the post-vaccine infection (PVI) group. P values determined using the Mann-Whitney U test (p = 0.0098). **(B)** Percentages of uninfected vs PVI participants depending on the anti-S IgG serum levels determined using the research assay. **(C)** Comparison of anti-S (WT) IgA serum levels between the uninfected group and the PVI group. P values determined using the Mann-Whitney U test (p = 0.1167). **(D)** Correlation between research assay anti-S (WT) IgG serum levels (BAU/ml) and Roche Elecsys® Anti-SARS-CoV-2 S assay anti-RBD (WT) total bAb serum levels (BAU/ml) in 171 samples (Spearman ρ = 0.8239; p < 0.0001). **(E)** Comparison of anti-RBD (WT) total bAb serum levels between the uninfected group and the PVI group using the Roche Elecsys® Anti-SARS-CoV-2 S assay. P values determined using the Mann-Whitney U test (p = 0.2138). **(F)** Correlation between research assay anti-S (WT) IgG serum levels (BAU/ml) and Siemens ADVIA Centaur® sCOVG assay anti-RBD (WT) IgG serum levels (BAU/ml) in 171 samples (Spearman ρ = 0.6929; p < 0.0001). **(G)** Comparison of anti-RBD (WT) IgG serum levels between the uninfected group and the PVI group using the Siemens ADVIA Centaur® sCOVG assay. P values determined using the Mann-Whitney U test (p = 0.4936). Dots indicate results from individual participants and bars indicate geometric mean with 95% confidence intervals (CI) in A, C, E, and G. Serum samples available for analysis: PVI group, n=31; uninfected group, n=140. * p < 0.05; ** p < 0.01; *** p < 0.001; **** p < 0.0001.

To evaluate the utility of FDA EUA commercial assays for anti-S antibodies, we tested all pre-Omicron wave serum samples for anti-RBD (WT) bAb levels using two manufacturer’s assays: Elecsys® Anti-SARS-CoV-2 S for cobas e analyzer (Roche, Basel, Switzerland) and ADVIA Centaur® sCOVG (Siemens, Erlangen, Germany). The maximum upper range for these assays as provided is 5,144 BAU/ml (after manufacturer recommended 1:20 dilution) and 1,781 BAU/ml, respectively. For the Roche assay we also ran 1:200 dilutions to enable a maximum upper range of 51,440 BAU/ml.

Strong correlations were observed between both the Roche and Siemens semi-quantitative commercial assays and the research MMIA for bAb to WT S (ρ=0.8239, p<0.0001 and ρ=0.6929, p<0.0001, respectively) **(Figures 1D, 1F)**. We diluted samples for the Roche assay 10-fold more than the original dilution to enable measurement of higher bAb concentrations. While anti-RBD (WT) total bAb serum levels were numerically greater in uninfected individuals compared to those with PVI (GM 13,819 vs 10,397), this difference did not reach statistical significance (p=0.2138) **(Figure 1E)**. A plot of incidence against antibody levels demonstrates that the percentage of PVIs reaches zero at value greater than 50,000 BAU/ml **(Supplementary Figure 2C)** when considering the Roche assay. Analyses of anti-RBD (WT) IgG levels obtained using the Siemens assay demonstrate no significant difference in bAb levels between the 2 groups, likely because we did not conduct additional dilutions and thus the majority of samples reached the maximum value measurable of the assay **(Figure 1G)**.

Finally, we also measured serum levels of IgG bAb against the Omicron subvariant BA.1 S protein. A strong correlation was observed between anti-S (BA.1) and anti-S (WT) IgG bAb levels (ρ=0.8411, p<0.0001) **(Figure 2A)**. Individuals that did not develop PVI during the initial Omicron wave had higher pre-exposures anti-S (BA.1) bAb levels than those that developed PVI (GM 276.9 AU/ml vs 179.9 AU/ml, p=0.04) **(Figure 2B)**.

**Figure 2.**
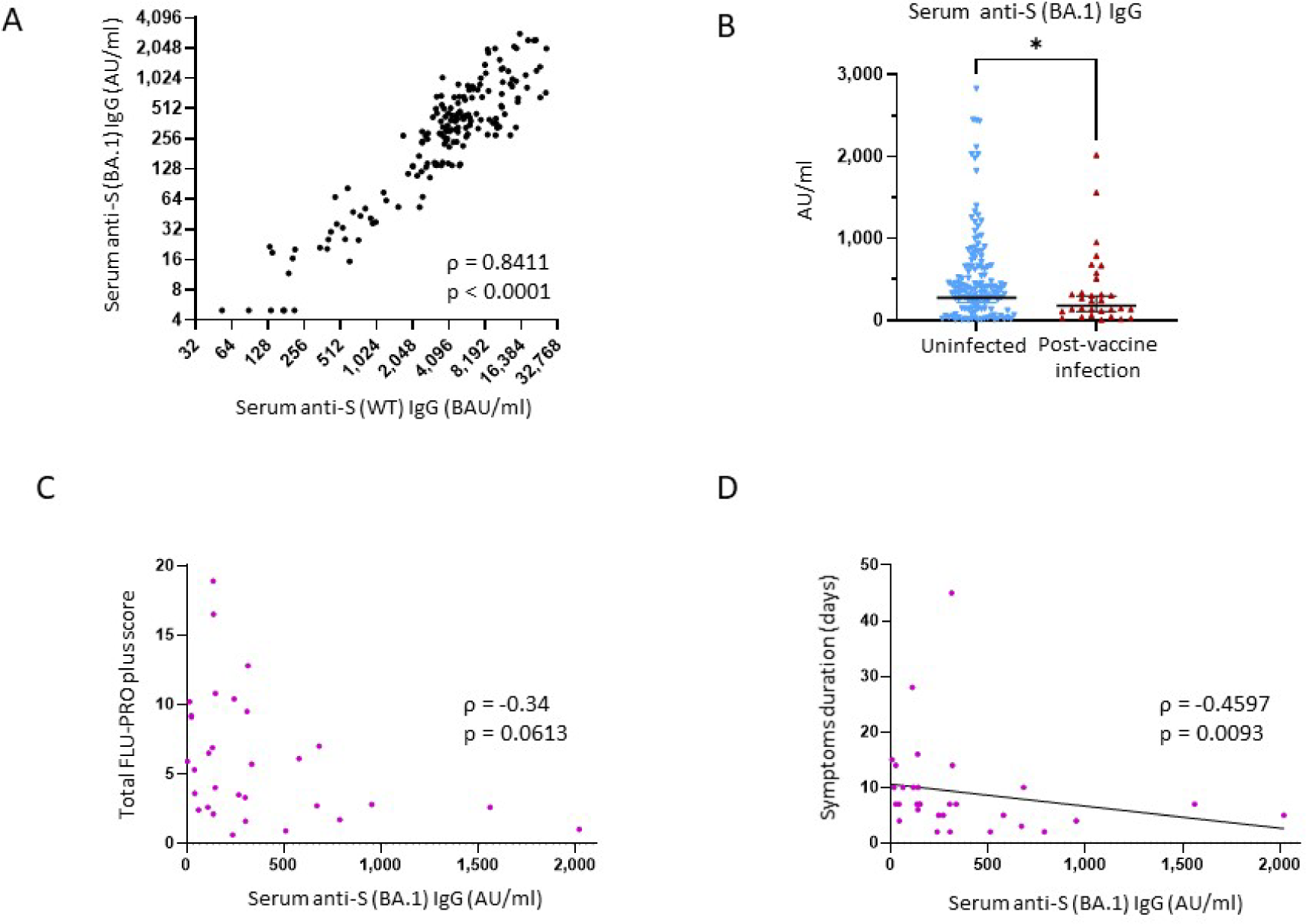
Binding antibody serum levels against Omicron subvariant BA.1 Spike in research assay and correlation with protection against SARS-CoV-2 post-vaccine infection. **(A)** Correlation between research assay anti-S (WT) IgG serum levels (BAU/ml) and research assay anti-S (BA.1) IgG serum levels (AU/ml) in 171 samples (Spearman ρ = 0.8411; p < 0.0001). **(B)** Comparison of anti-S (BA.1) IgG serum levels between the uninfected group and the PVI group. P values determined using the Mann-Whitney U test (p = 0.04). **(C)** Correlation between research assay anti-S (BA.1) IgG serum levels (AU/ml) and total FLU-PRO plus symptom scores (n=31; Spearman ρ = −0.34; p = 0.0613). **(D)** Correlation between research assay anti-S (BA.1) IgG serum levels (AU/ml) and total symptom duration of any symptom in days (n=31; Spearman ρ = −0.4597; p = 0.0093). Dots indicate results from individual participants. * p < 0.05; ** p < 0.01; *** p < 0.001; **** p < 0.0001.

### 3.4. Pre-Omicron wave binding IgG serum levels correlate inversely with symptom severity and duration

All 32 individuals that developed PVI during the observation period had a symptomatic, outpatient disease presentation. Assessment of symptoms by FLU-PRO plus score demonstrates a wide range of symptom severity, with a range of scores from 0.6 to 18.9 out of a maximum possible score of 28. Symptoms in the nasal, throat, body/systemic, chest, and sense categories were the most severe **(Figure 3A)**. Median duration of symptoms was 7 days (range 2 – 45), with 12 of 32 (37.5%) of individuals reporting symptoms lasting for 10 days or more.

**Figure 3.**
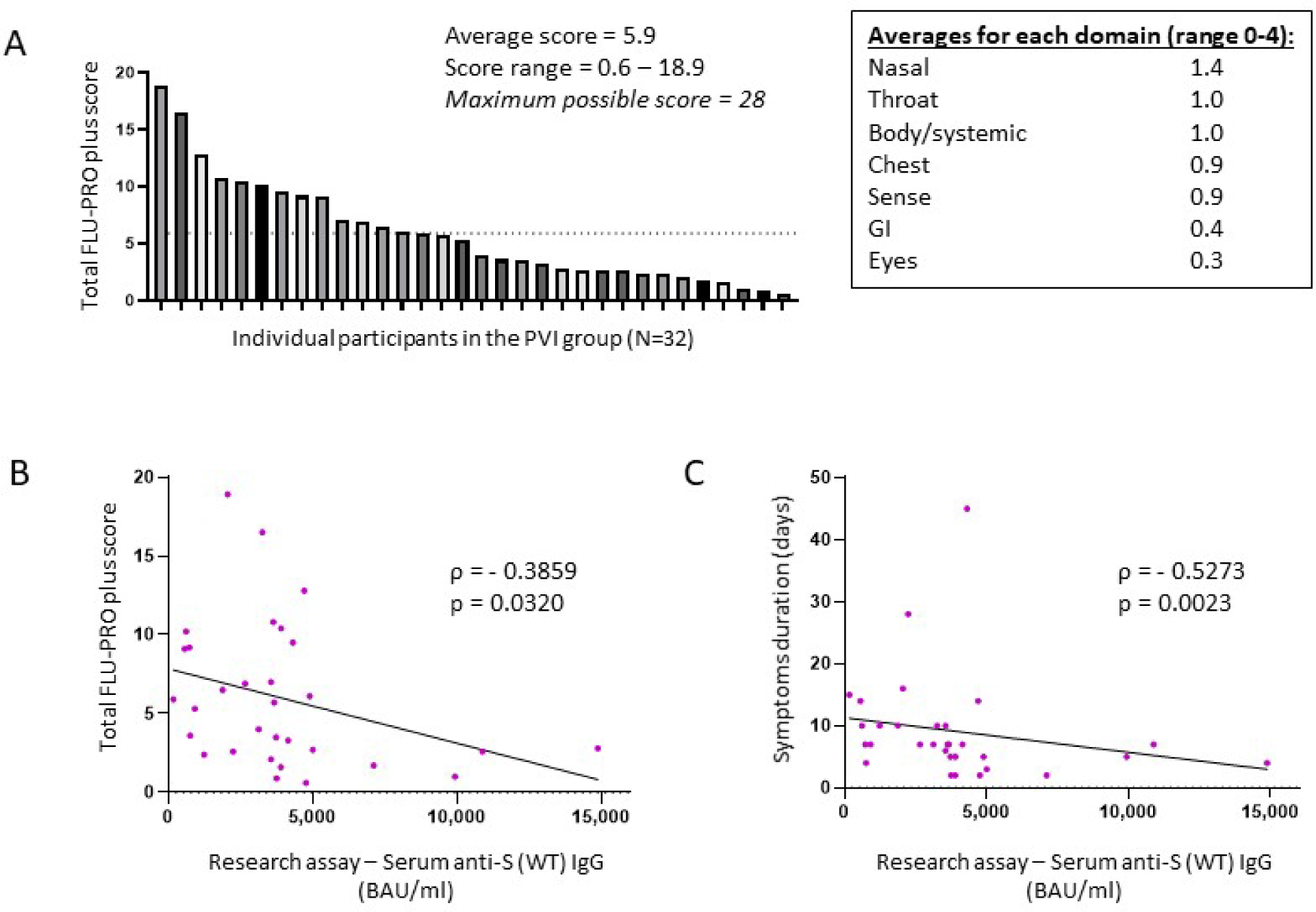
Total FLU-PRO plus symptom scores and average scores for each symptom domain in the post-vaccine infection group. **(A)** Total FLU-PRO plus symptom scores for each participant in the PVI group (n=32), and average scores for each symptom domain. The dashed line represents the average score of 5.9. **(B)** Correlation between research assay anti-S (WT) IgG serum levels (BAU/ml) and total FLU-PRO plus symptom scores (n=31; Spearman ρ = −0.3859; p = 0.032). **(C)** Correlation between research assay anti-S (WT) IgG serum levels (BAU/ml) and total symptom duration of any symptom in days (n=31; Spearman ρ = −0.5273; p = 0.0023). Dots indicate results from individual participants. * p < 0.05; ** p < 0.01; *** p < 0.001; **** p < 0.0001.

Higher anti-S (WT) IgG bAb serum levels correlated with a reduced symptom severity (ρ= - 0.3859, p=0.032) **(Figure 3B)** and with reduced duration of symptoms (ρ= −0.5273, p=0.0023) **(Figure 3C)**. When analyzed per symptom domain, higher IgG bAb serum levels correlated with reduced symptom severity in the nasal (ρ= −0.4654, p=0.0083), eyes (ρ= −0.4145, p=0.0204) and body/systemic domains (ρ= −0.4902, p=0.0051), but not in the throat (ρ= −0.1471, p=0.4299), chest (ρ=0.01075, p=0.9542), gastrointestinal (ρ= −0.3224, p=0.0769), and smell/taste domains (ρ= −0.04742, p=0.8) **(Supplementary Figure 3)**. As with anti-S (WT) IgG bAb, high anti-S (BA.1) IgG bAb serum levels were also correlated with decreased symptom duration (ρ= −0.4597, p=0.0093) **(Figure 2D)**. Anti-S (BA.1) IgG bAb levels also exhibited a correlation with reduced symptom severity scores, but the relationship did not reach statistical significance (ρ= −0.34, p=0.0613) **(Figure 2C)**.

### 3.5. Pre-Omicron wave binding IgG and IgA salivary levels did not correlate with protection against infection

Saliva samples were obtained during the Fall 2021 research clinic visit, at a median of 52.5 (IQR: 45.8 – 67.3) days after last immunization for the PVI group (participants, N=32; saliva samples available for analysis, n=32) and 46.5 (IQR: 32.8 – 58.3) days after last immunization for the uninfected group (participants, N=144; saliva samples available for analysis, n=143).

No significant differences were found when comparing the saliva levels of anti-S (WT) IgG, anti-S (WT) IgA, and anti-S (WT) sIgA bAbs between the uninfected and the PVI groups **(Figures 4A, 4B, Supplementary Figure 4)**. When looking at the distribution of anti-S IgG bAb salivary levels, the percentage of PVIs among the study participants slightly decreased from 23.1% and 25.0% for levels of ≤55 AU/ml and 55-250 AU/ml, respectively, to 15.3% at levels of 250-1,000 AU/ml, and 16.7% at ≥1,000 AU/ml **(Figure 4C)**. When looking at the distribution of anti-S (WT) IgA bAb salivary levels, the percentage of PVIs among the study participants remain stable from 19.3% and 14.3% for levels of ≤55 AU/ml and 55-100 AU/ml, respectively, to 16.7% at 100-200 AU/ml, and 20.0% at ≥200 AU/ml **(Figure 4D)**. There was a robust correlation between the saliva and serum levels observed for both IgG and IgA bAbs, with a Spearman’s rank correlation coefficient of ρ=0.5536, p<0.0001 for anti-S (WT) IgG bAb levels **(Figure 4E)**, and a Spearman’s rank correlation coefficient of ρ=0.3406, p<0.0001 for anti-S (WT) IgA bAb levels in serum and saliva **(Figure 4F)**. One reason why saliva bAbs in this cohort study may not have correlated with protection in this study was because only a few individuals had hybrid immunity and because mRNA vaccination alone induced relatively low levels of saliva antibody levels.

**Figure 4.**
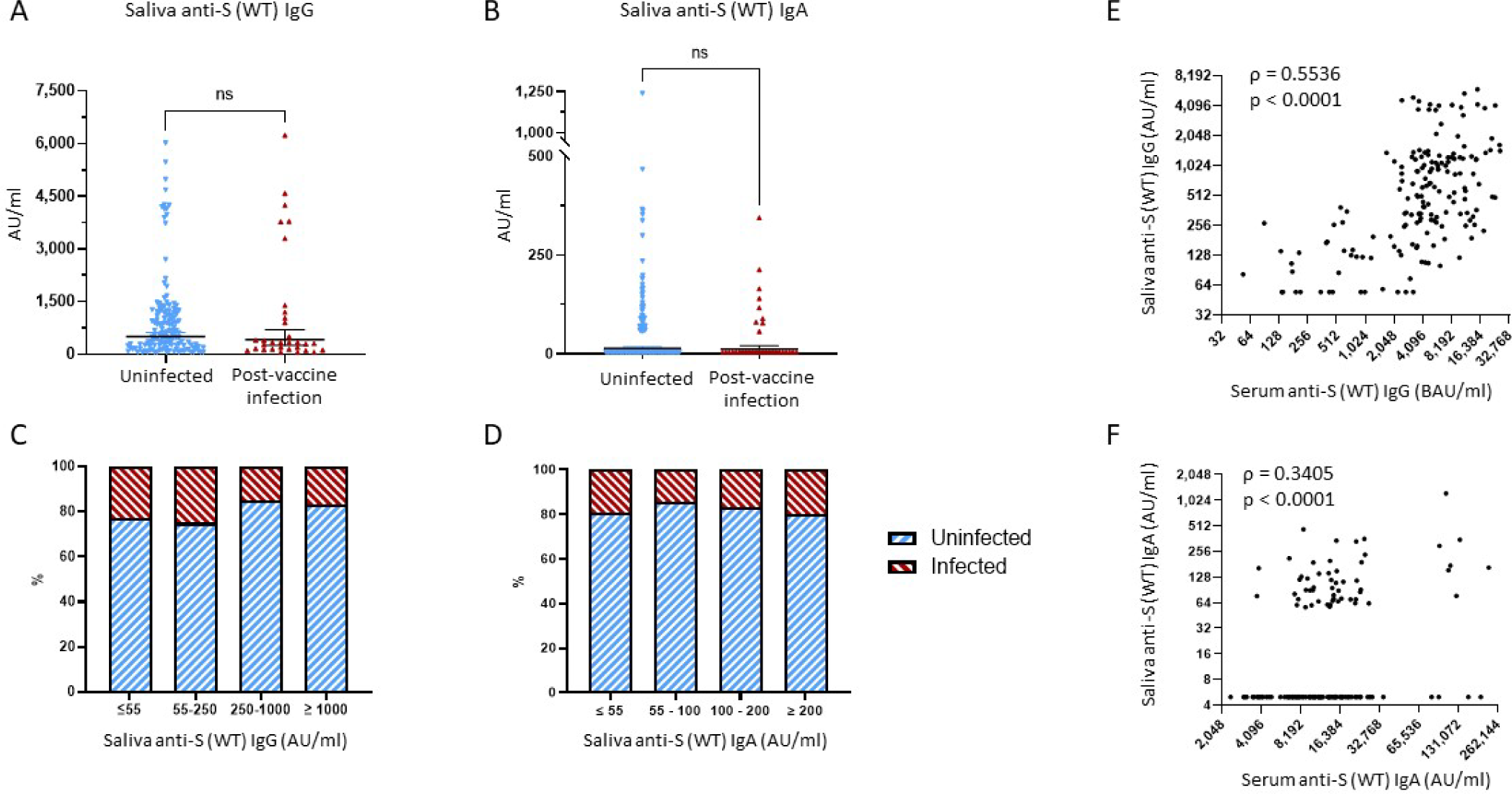
Binding antibody saliva levels against WT Spike. **(A)** Comparison of anti-S (WT) IgG saliva levels between the uninfected group and the PVI group. P values determined using the Mann-Whitney U test (p = 0.2786). **(B)** Comparison of anti-S (WT) IgA saliva levels between the uninfected group and the PVI group. P values determined using the Mann-Whitney U test (p = 0.6579). **(C)** Percentages of uninfected vs PVI participants depending on the anti-S (WT) IgG saliva levels. **(D)** Percentages of uninfected vs PVI participants depending on the anti-S (WT) IgA saliva levels. **(E)** Correlation between research assay anti-S (WT) IgG serum levels (BAU/ml) and anti-S (WT) IgG saliva levels (AU/ml) (n=175; Spearman ρ = 0.5536; p < 0.0001). **(F)** Correlation between research assay anti-S (WT) IgA serum levels (AU/ml) and anti-S (WT) IgA saliva levels (AU/ml) (n=175; Spearman ρ = 0.3405; p < 0.0001). Dots indicate results from individual participants and bars indicate geometric mean with 95% confidence intervals (CI) in A and B. Saliva samples available for analysis: PVI group, n=32; uninfected group, n=144. * p < 0.05; ** p < 0.01; *** p < 0.001; **** p < 0.0001.

### 3.6. Neutralization titers against Omicron variants correlate with protection against post-vaccine infections

We compared the NTs of the 171 serum samples collected during the Fall 2021 research clinic visit against pseudoviruses bearing spike proteins from 4 variants: D614G, Delta (B.1.617.2), and 2 Omicron subvariants (BA.1, BA.1.1).

No significant differences were observed between uninfected and PVI groups when considering D614G and Delta (p=0.1443, and p=0.0894, respectively). NTs against D614G were 1.45-fold lower in the PVI group (geometric mean titer [GMT]=2,674.0) than the uninfected group (GMT=3,883.0), and NTs against Delta were 1.49-fold lower in the PVI group (GMT=972.0) than the uninfected group (GMT=1,450.0), but these differences did not reach statistical significance **(Figure 5)**. When considering the two Omicron subvariants BA.1 and BA.1.1, a significant difference was observed between NTs in the PVI and uninfected groups (p=0.031, and p=0.021, respectively). NTs against BA.1 were 1.72-fold lower in the PVI group (GMT=286.2) than the uninfected group (GMT=493.6), and NTs against BA.1.1 were 1.82-fold lower in the PVI group (GMT=302.5) than the uninfected group (GMT=552.0) **(Figure 5)**. When plotting neutralization titers against the incidence of infection in individuals above that level, the percentage of PVIs reach zero at an ID_50_ value equal or greater than 20,000 for D614G, 7,000 for Delta B.1.617.2, 2,700 for Omicron BA.1, and 2,700 for Omicron BA.1.1 **(Supplementary Figures 2E, 2F, 2G, 2H**).

**Figure 5.**
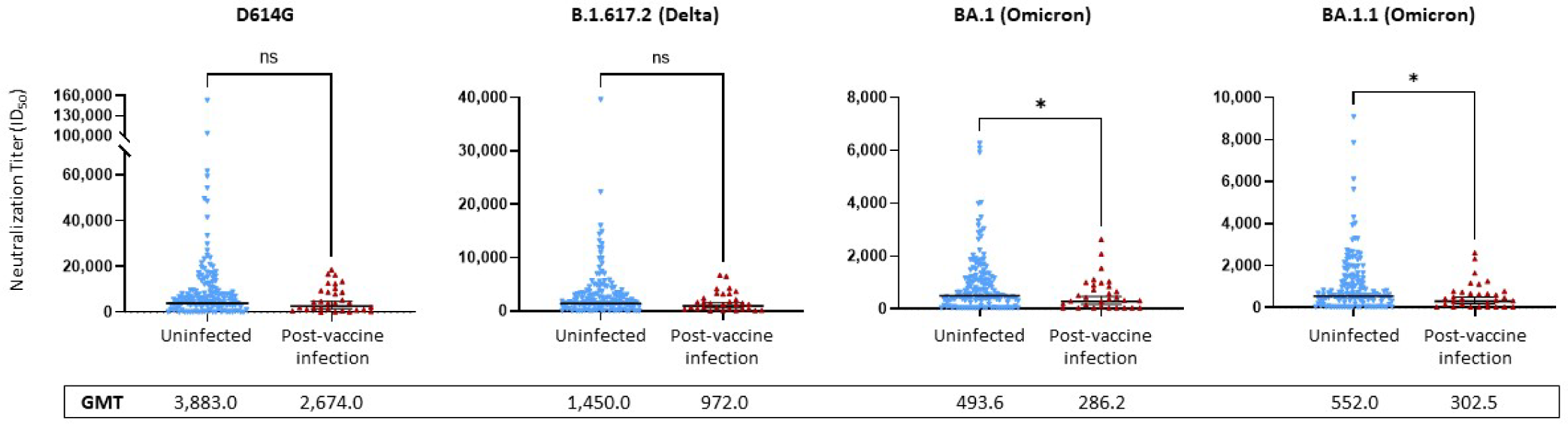
Pseudovirus neutralization ID_50_ titers against D614G, Delta variant B.1.617.2, and Omicron subvariants BA.1 and BA.1.1. Neutralization assays used lentiviral pseudoviruses bearing SARS-CoV-2 spike proteins from D614G, Delta variant B.1.617.2, or Omicron subvariants BA.1 and BA.1.1. P values determined using the Mann-Whitney U test (D614G, p = 0.1443; B.1.617.2, p = 0.0894; BA.1, p = 0.0313; BA.1.1, p = 0.021). Dots indicate results from individual participants and bars represent geometric mean titers (GMT) with 95% confidence intervals (CI), and GMTs are indicated. Serum samples available for analysis: PVI group, n=31; uninfected group, n=140. * p < 0.05; ** p < 0.01; *** p < 0.001; **** p < 0.0001.

When comparing anti-S IgG bAbs to NTs, we found strong correlations between anti-S (WT) IgG bAb serum levels and NTs against D614G (ρ=0.7801, p<0.0001) **(Supplementary Figure 5A**), between anti-S (WT) IgG bAb serum levels and NTs against Omicron BA.1 (ρ=0.7889, p<0.0001) **(Supplementary Figure 5B)**, between anti-S (BA.1) IgG bAb serum levels and NTs against D614G (ρ=0.7963, p<0.0001) **(Supplementary Figure 5C)**, and between anti-S (BA.1) IgG bAb serum levels and NTs against Omicron BA.1 (ρ=0.8145, p<0.0001) **(Supplementary Figure 5D)**.

### 3.7. Home risk score correlates strongly with risk of post-vaccine infection

Questionnaires on risk exposures and precautionary behaviors were completed by participants during the Spring 2022 visit (first visit after April 1 2022) and covered the preceding time period since the Fall 2021 visit. While there was a broad range of potential work-related exposures in the cohort (median score of 66.7, IQR: 33.3 – 66.7, for both study groups), most individuals practiced high degrees of precautionary measures while at work (median score of 91.7, IQR: 83.3 – 100.0, for both study groups). Neither work risk scores (WRS) nor work precautionary scores (WPS) were significantly different between uninfected and PVI groups. While home precautionary scores (HPS) exhibited substantial variability between study participants (median score of 46.5, IQR: 36.0 – 56.0 and median score of 49.5, IQR: 27.2 – 61.0 for uninfected vs PVI group, respectively), no difference was observed in scores between those that did and did not develop PVIs **(Figure 6A)**.

**Figure 6.**
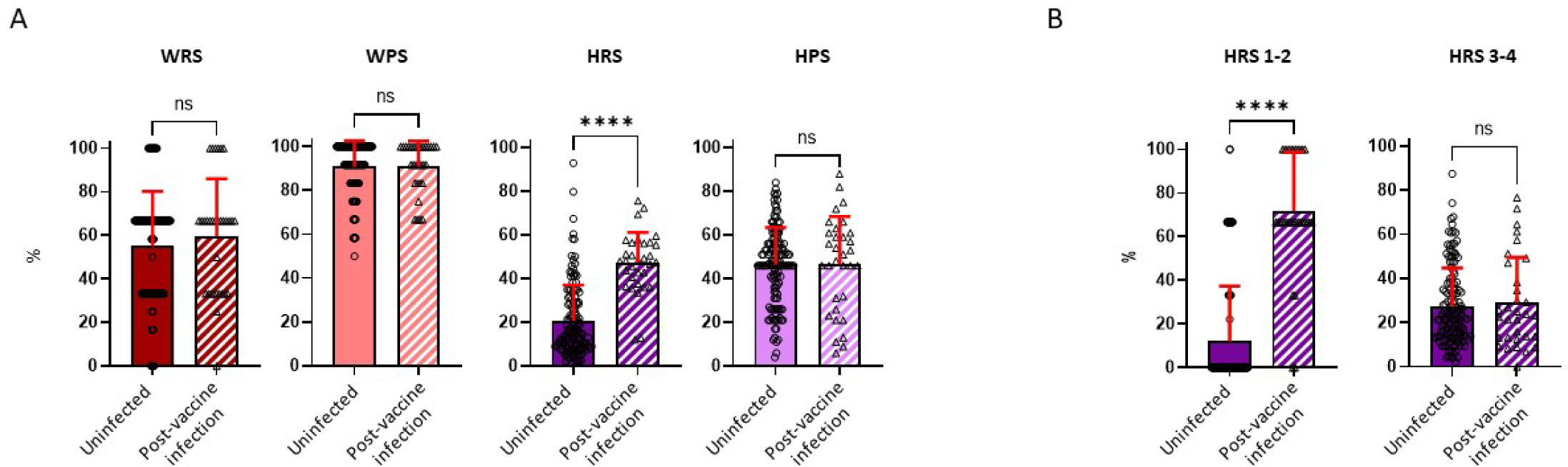
Risk exposure and precautionary measure scores calculated for the workplace and home. Risk exposure and precautionary measure scores were obtained during the visit following the Fall 2021 research clinic visit (January to March 2022). **(A)** Work risk score (WRS), calculated from 3 questions (uninfected, n=140; PVI, n=32); Work precautionary score (WPS), calculated from 3 questions (uninfected, n=134; PVI, n=31); Home risk score (HRS), calculated from 4 questions (uninfected, n=140; PVI, n=32); Home precautionary score (HPS), calculated from 5 questions (uninfected, n=140; PVI, n=32). **(B)** HRS 1-2 represents questions 1 and 2 of the HRS category (COVID-19 exposure in household). HRS 3-4 represents questions 3 and 4 of the HRS category (out-of-the-house activities and social gatherings). P values determined using the Mann-Whitney U test (WRS, p=0.3797; WPS, p=0.9745; HRS, p<0.0001; HPS, p=0.6852; HRS 1-2, p<0.0001; HRS 3-4, p=0.9228). Dots indicate results from individual participants and bars indicate mean with standard deviation. * p < 0.05; ** p < 0.01; *** p < 0.001; **** p < 0.0001.

In contrast to WRS, WPS, and HPS, home risk scores (HRS) were significantly increased in individuals that developed PVI. The mean HRS was 2.3-fold higher in the PVI group (mean=47.33) than in the uninfected group (mean=20.77, p<0.0001) **(Figure 6A)**. To ascertain if there was a specific component of the HRS questionnaire that was influencing risk, we divided the HRS questionnaire into 2 main subcategories: HRS questions 1 and 2 that related to whether others in the household had COVID-19, and HRS questions 3-4 which asked about extent of out-of-house activities and social gatherings **(Supplementary Table 1)**. The main determinant of COVID-19 risk was whether another individual in the household had (or potentially had) COVID-19, as scores for HRS questions 1-2 were 5.9-fold higher in individuals with PVI than in those that were uninfected (p < 0.0001) whereas there were no significant differences in scores for HRS questions 3-4 **(Figure 6B)**.

### 3.8. Pre-Omicron wave serum anti-S IgG binding antibody level remains a strong correlate of protection against post-vaccine infections when adjusting for demography and risk behavior

Univariate logistic regression analyses showed that being female (OR=0.406, p=0.025), serum level of anti-S (WT) IgG bAb (OR=0.703, p=0.016), and neutralization titers against the Omicron subvariant BA.1.1 (OR=0.573, p=0.035) significantly correlated with protection against PVIs, while the home risk score correlated with increased risk of PVI (OR=2.144, p<0.0001). While not reaching statistical significance, NTs against Delta B.1.617.2 (OR=0.498, p=0.067) and Omicron BA.1 (OR=0.694, p=0.056) were associated with a reduced risk of PVI **(Figure 7A, Supplementary Table 3)**. We then conducted multivariable logistic regression analysis for each immunological variable with adjustment for the non-immunological factors of age, sex, and home risk score. After controlling for these non-immunological factors, only anti-S (WT) IgG bAb, measured by the research MMIA, remained significantly correlated with protection against PVI (OR=0.673, p=0.025). Levels of IgG bAb to BA.1 S protein (OR=0.811, p=0.134) performed less well than IgG bAb to WT S protein, even though cases were predominantly due to Omicron variants. NTs to BA.1.1 were also strongly associated with protection against PVI (OR 0.511), though the p value was not statistically significant (p=0.056) **(Figure 7B, Supplementary Table 3)**. Of note, pseudoR2 value for age, sex, home risk score, and bAb level was 0.375, suggesting that the final model fits 37.5% better than a model that predicts the same risk of acquiring SARS-CoV-2 infection for all individuals.

**Figure 7.**
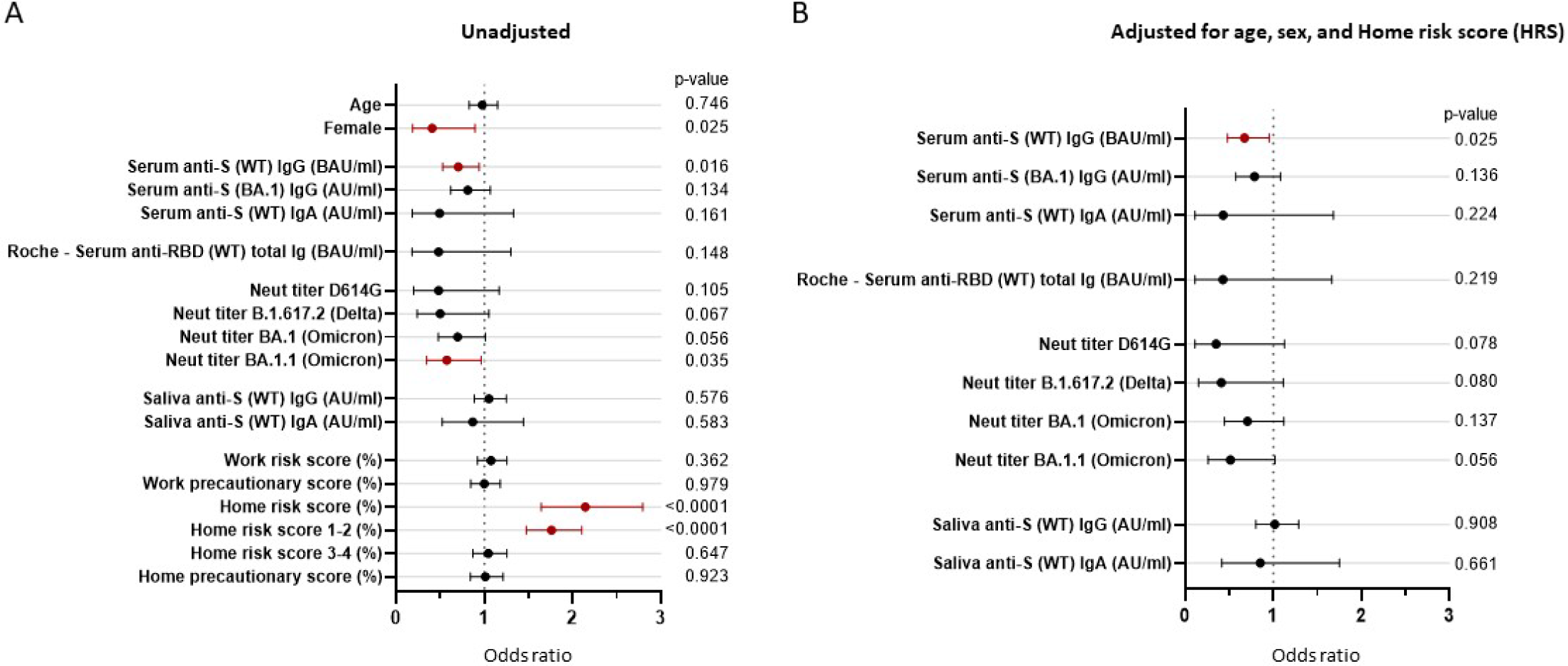
Odds ratios unadjusted and adjusted for non-immunological factors. Odds ratios (OR) represent the relative change in the odds of post-vaccine infection corresponding to an increase in the independent variable of 1/10 of its range (except for the odds ratio for “Female”). **(A)** Unadjusted odds ratios. **(B)** The pre-Omicron wave covariates were adjusted for age, sex and home risk score (non-immunological factors). CI, confidence interval; CI, confidence intervals; ID_50_, 50% inhibitory dilution; BAU, binding antibody units; AU, arbitrary units.

## 4. Discussion

Identifying biomarkers that can serve as simple and reproducible correlates of subsequent immunity for SARS-CoV-2 infection and associated COVID-19 disease has been a goal of the scientific community since the onset of the pandemic. In addition to potentially providing mechanistic insights into host defense against SARS-CoV-2, correlates of protection can be used for informing recommendations on frequencies of booster vaccinations and booster compositions, assessing potential efficacy of novel vaccines, and guiding patient-specific interventions in immunocompromised and other high-risk individuals.

In this study, we evaluated multiple serological and salivary markers for correlation with protection against symptomatic SARS-CoV-2 infection in a cohort of generally healthy, vaccinated, adult healthcare workers. We used a multivariable regression approach that incorporates patient demography and risk behavior into immune correlate of protection estimates and minimized bias; other COVID-19 vaccine correlate of protection estimates which do not include these variables may be subject to confounding. Other strengths of the study include obtainment of serum just before the initial Omicron wave in the U.S., use of a generally healthy adult population that all work at the same healthcare center, evaluation and control for individual risk exposures and mitigation behaviors, assessments for severity and duration of infection using a validated symptom questionnaire, use of WT and Omicron-specific NTs, and bAb assays, and comparisons between a research-based bAb assay with commercially available assays. Key immunological findings include: 1) research assay anti-S IgG bAb levels (directed against either WT or Omicron BA.1) NTs against Omicron variants correlated strongly with protection against symptomatic infection, 2) higher anti-S IgG bAb levels correlated inversely with symptom severity and illness duration in individuals that developed post-vaccine infection (PVI), and 3) anti-S IgG bAb levels greater than 5,000 BAU/ml are required to achieve robust protection against symptomatic infection with Omicron variants (**Figure 1B**). Further, as discussed in more detail below, results of this study also suggest that commercial assays for anti-RBD bAb levels may need to be reformatted to enable detection of higher maximum values for use as predictors of reduced susceptibility to SARS-CoV-2 infection. With regards to behavioral analyses, we found that home risk score, driven primarily by household SARS-CoV-2 exposure, strongly correlated with increased risk of SARS-CoV-2 infection. In contrast, we observed no correlation between risk of infection and measures of work exposure risk, work precautionary behaviors, and home (outside of work) precautionary measures.

The finding that both anti-S bAb and NTs correlate strongly with protection against SARS-CoV-2 infection is consistent with a number of studies that have evaluated this issue previously ^2,4,5,8,11–17^. Interestingly, even though NTs are often the principal biomarkers used to inform policy recommendations on booster doses ^30,31^, our findings suggest that bAb levels may have similar utility for this purpose. In unadjusted analyses, both bAb (to either WT or Omicron) and NTs (to Omicron BA.1.1) were significantly lower in individuals that developed a PVI. After regression analysis adjusting for age, sex, and home SARS-CoV-2 risk score, only bAb levels retained statistical significance (p value of 0.025) with the p value for NTs to BA.1.1 increasing to 0.056. The finding that bAb levels correlate as well or better than NTs has been observed in a number of other studies ^11,12,14^. Interestingly, a study that incorporated data from 7 clinical studies of four different vaccine platforms found that IgG bAb levels induced by the different vaccines correlated better than NTs in explaining differences in vaccine efficacy ^13^.

A number of factors may account for why bAb levels function at least as well as NTs as correlates of immunity for subsequent SARS-CoV-2 infection. First, while inhibiting viral entry is a key mechanism by which antibodies protect against SARS-CoV-2, it is likely that other effector functions of antibodies also play a role in protection which are not accounted for when assessing neutralizing activity. Second, it is possible that NT assays are simply more variable and less precise than bAb assays. When measured against a pooled convalescent standard, results from different NT assays varied by as much as 100-fold ^32^. Additionally, in contrast to NTs, which typically provide outcomes in two-fold increments, bAbs can provide quantitative results in continuous increments.

Notably, the bAb levels that correlated with substantial protection were markedly higher in our study than in previous reports. Whereas earlier studies evaluating correlates of protection for SARS-CoV-2 often found that anti-S IgG bAb levels > 250 or 1,000 BAU/ml afforded substantial protection against PVI ^2,14,17^, we observed that BAU levels greater than 5,000 correlated with > 90% protection against PVI during the early Omicron era. This may be because our study evaluated for PVIs during the peak of SARS-CoV-2 transmission intensity in the U.S., because Omicron variants are highly infectious, and because most Ab were not Omicron-specific. Consistent with our findings, a recent study in Japan observed that 43,000 AU/ml (approximately 6,000 BAU/ml) measured by the Abbott AdviseDx SARS-CoV-2 IgG II assay (Abbott, Sligo, Ireland) correlated with 80% protection in infection-naïve individuals during the Omicron BA1.5 wave in Japan ^33^.

With regards to variant-specific bAb, our study found that bAb levels to WT S protein correlated with protection as well as bAb to Omicron BA.1 S protein for protection. Because individuals in this study had not yet received bivalent mRNA vaccines, and only a few had hybrid immunity, the quantitative level of bAbs observed to WT may reflect the general strength of all adaptive immune responses developed against the vaccine. In contrast, measuring the level of bAb against a non-vaccine variant spike protein would primarily measure cross-protective Abs.

Another key finding in our study is that higher bAb levels correlated inversely with symptom severity and duration of illness in those individuals that developed PVIs. While none of the participants in our study required supplemental oxygen or hospitalization, 34% had symptoms that persisted for more than one week. Two other studies have found that high titers of anti-S bAb at time of diagnosis and admission for COVID are associated with less progression to severe disease ^7,34^, and, consistent with this study, one other study observed that pre-infection bAb and NT levels correlate with reduced symptom severity ^35^. The observation that higher bAb levels are associated with less severe illness provides a potential rationale on an individual basis for providing booster vaccinations to generally healthy adults immediately prior to anticipated peak COVID season.

We also analyzed salivary S-specific IgG and IgA bAb levels. While no significant association was observed between the levels of these antibodies and development of PVI, this does not mean that high levels of salivary antibodies would not correlate with protection. We speculate that salivary antibodies did not serve as correlates of immunity because their levels were relatively low due to weak induction of mucosal antibodies by the intramuscular BNT162b2 mRNA vaccine.

In addition to conducting serological studies, we also obtained information on work and home risk exposures and precautionary measures. Of these, only home risk score was significantly associated with risk for SARS-CoV-2 infection, and this risk was driven predominantly by whether individuals had a household member with recent COVID. Interestingly, while study participants had a wide range of risk exposure scores to SARS-CoV-2 while working in the hospital, work risk exposure scores did not correlate with likelihood of getting infected. We suspect this is due to the strict adherence to precautionary measures (reflected by the very high work precautionary scores) exhibited by most participants while in the workplace. Of note, we also did not observe a correlation between home precautionary scores, which included stringency of mask wearing and type of mask worn, and risk of COVID infection. Recent systematic reviews have concluded that mask wearing is associated with, at most, small decreases in risk for SARS-CoV-2 infection in the community setting ^36,37^. We suspect our sample size may have been too small to observe a benefit from community mask wearing.

Finally, we also compared our bAb assay with those of two commercial systems, the Roche (Elecsys® anti-SARS-CoV-2 S assay) and the Siemens (ADVIA Centaur® SARS-CoV-2 IgG). While both assays showed strong correlation with our in-house research assay (with the Roche assay providing values moderately higher than the research assay and the Siemens assay providing values moderately lower than the research assay), neither reached statistical significance when evaluating for differences in levels between participants with and without PVI. One possible explanation for why the research bAb assay functioned better as a correlate of protection is that it assessed for antibodies against full pre-fusion stabilized ectodomain trimers of the spike protein, whereas the commercial assays only measured anti-RBD antibodies. Since antibodies to non-RBD regions can exert antiviral effects via modalities such as complement activation, opsonophagocytosis, and antibody-dependent cellular cytotoxicity, it is possible that the research assay better captured the full spectrum of antibody immunity against SARS-CoV-2. We suspect, however, that the commercial assays may not have performed quite as well as the research assay because they were not designed to accurately measure high bAb levels. Our in-house research bAb test is a microsphere assay based on Luminex technology that has a large dynamic range, likely enabling accurate measurement of bAb levels even at high concentrations. In contrast, the Roche assay was designed to detect a maximum bAb level of 5,144 BAU/ml and the Siemens a maximum range of 1,781 BAU/ml. In our study, we conducted a second, non-standard dilution of serum samples when running samples on the Roche system, but this may have increased the variance observed at high concentrations (reflected by the greater spread of values compared to our research assay at the high end of BAUs in **Figure 1D**). In studies using much larger cohorts, bAb levels measured by the Roche Elecsys assay were found to significantly correlate with infection risk ^5,17^. Given that we observed > 90% protection at BAUs greater than 5,000 BAU/ml, the results of this study suggest that commercial assays could be redesigned with a higher top range than currently available.

The key limitations of this study are the moderate sample size and the lack of T cell assays. Recent studies have observed that SARS-CoV-2-specific T cell responses correlate with protection against severity of clinical disease ^38–40^. We were not able to conduct T cell studies in this analysis because the timepoint just prior to the Omicron variant epidemic was not one at which PBMCs were drawn in the PASS study. Future studies should directly compare how well B and T cell responses function as independent and synergistic correlates of immunity, ideally incorporating demographic and risk behavior data in the multivariate regression framework demonstrated here. Additionally, as the study was of generally healthy younger adults, the results may not be generalizable to all populations, especially children, older adults, or immunocompromised individuals in whom other biomarkers may be more suitable correlates of immunity, particularly for endpoints such as hospitalization ^41^. Finally, the sample evaluated was restricted to a discrete era (the first U.S. Omicron wave), and thus the results may not be generalizable to current and future eras characterized by emerging new variants and increasingly complex population SARS-CoV-2 immunity.

Finally, it is important to note that while we identified independent immunological (bAb levels) and behavioral (household exposure risk) predictors of infection, no single factor served as a highly accurate correlate of protection. Indeed, the pseudoR2 analysis we conducted suggests that a model that includes age, sex, home risk score, and bAb level is only 37.5% better at predicting who would have developed symptomatic infection than a model with no predictors. These limits call for an intensive, large scale study that prospectively assesses numerous factors associated with viral risk susceptibility, including innate immune traits, adaptive immune traits, known genetic risks, and lifestyle factors such as sleep and exercise, to be able to better construct a predictive model of resilience against SARS-CoV-2 infection.

## 6. Disclaimer

The views expressed in this presentation are the sole responsibility of the presenter and do not necessarily reflect the views, opinions, or policies of the Uniformed Services University of the Health Sciences, the Department of Defense, the Department of the Navy, Army, Air Force, the United States Government, or the Henry M. Jackson Foundation for the Advancement of Military Medicine, Inc. (HJF). Several of the authors are U.S. Government employees. This work was prepared as part of their official duties. Title 17 US.C. § 105 provides that “Copyright protection under this title is not available for any work of the United States Government.” Title 17 US.C. § 101 defines a U.S. Government work as a work prepared by a military service member or employee of the U.S. Government as part of that person’s official duties. Mention of trade names, commercial products, or organizations does not imply endorsement by the U.S. Government. The study protocol was approved by the USUHS Institutional Review Board in compliance with all applicable federal regulations governing the protection of human subjects.

## 7. Author Contributions

E.G. designed study protocol, processed biological samples, analyzed the data and wrote the manuscript. C.H.O. performed the statistical analysis, read and approved the final manuscript. W.A.M. read, edited, and approved the final manuscript. S.A. performed the commercial serum bAb assays. J.H.P. designed the FLU-PRO Plus symptom questionnaire, and read, edited, and approved the final manuscript. T.L.C. performed the saliva MMIA assays. S.A.C., E.C.S., M.S.T. performed the serum MMIA assays. W.W., R.W. performed the pseudovirus production and neutralization assays. L.I., M.S.E., B.M.J.T., A.R.L. designed study protocol. G.W., Y.A., M.A.W., B.B., P.K., L.K. scheduled and conducted research clinic visits and biological sample collection. M.H.P., D.S., R.M. set up research clinic visits. K.B.L., R.Z.C., C.E.A., L.J.V., M.F., A.E.L., F.M. performed the virus sequencing and bioinformatics assays. O.O., J.D. set up the database. E.P. read, edited, and approved the final manuscript. H.H.H., M.S.M. processed biological samples. E.K.G. designed and analyzed the risk exposure and precautionary measure questionnaires. A.M.W.M. read, edited, and approved the final manuscript. D.R.T. read, edited, and approved the final manuscript. T.H.B. read, edited, and approved the final manuscript. W.C., S.R. organized nasopharyngeal swab collection and storage for virus sequencing. C.C.B. read, and approved the final manuscript. R.J.O. read, edited, and approved the final manuscript. C.D.W. read, edited, and approved the final manuscript. S.P. read, edited, and approved the final manuscript. E.D.L. read, edited, and approved the final manuscript. E.M. designed the study, and read, edited, and approved the final manuscript.

## Acknowledgements

The investigators gratefully acknowledge all research participants for their many contributions to the PASS study.

