## Supplemental Material for "Immune and behavioral correlates of protection against symptomatic post-vaccination SARS-CoV-2 infection"

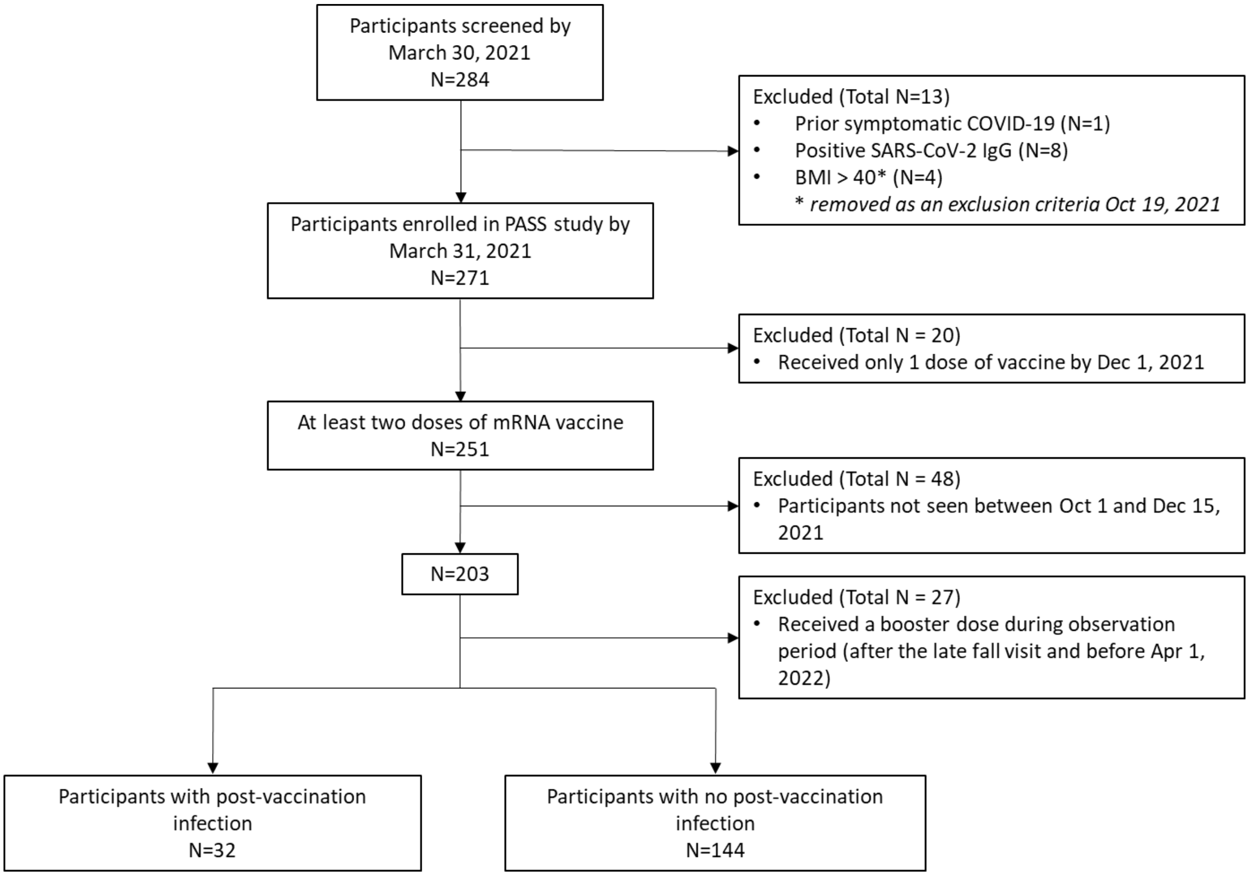


**Supplementary Figure 1. Strobe chart of the study cohort**

N represents the number of participants. BMI, Body Mass Index; PASS, Prospective Assessment of SARS-CoV-2 Seroconversion.

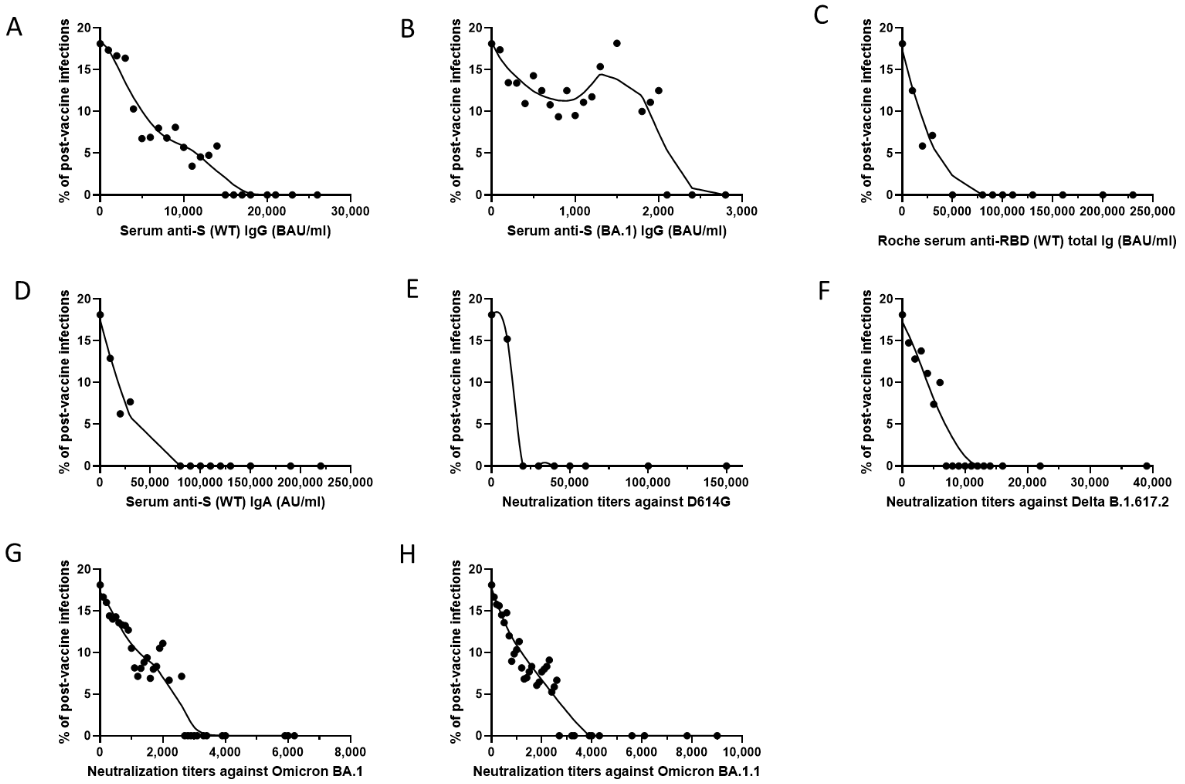


**Supplementary Figure 2. ROC curves – Probability of post-vaccine SARS-CoV-2 infection as a function of immune markers**

(A) Serum anti-S (WT) IgG measured by research assay. (B) Serum anti-S (BA.1) IgG measured by research assay. (C) Serum anti-RBD (WT) total Ig measured by Roche Elecsys® Anti-SARS-CoV-2 S assay. (D) Serum anti-S (WT) IgA measured by research assay. (E) Pseudovirus neutralization antibody titers against D614G. (F) Pseudovirus neutralization antibody titers against B.1.617.2. (G) Pseudovirus neutralization antibody titers against BA.1. (H) Pseudovirus neutralization antibody titers against BA.1.1.

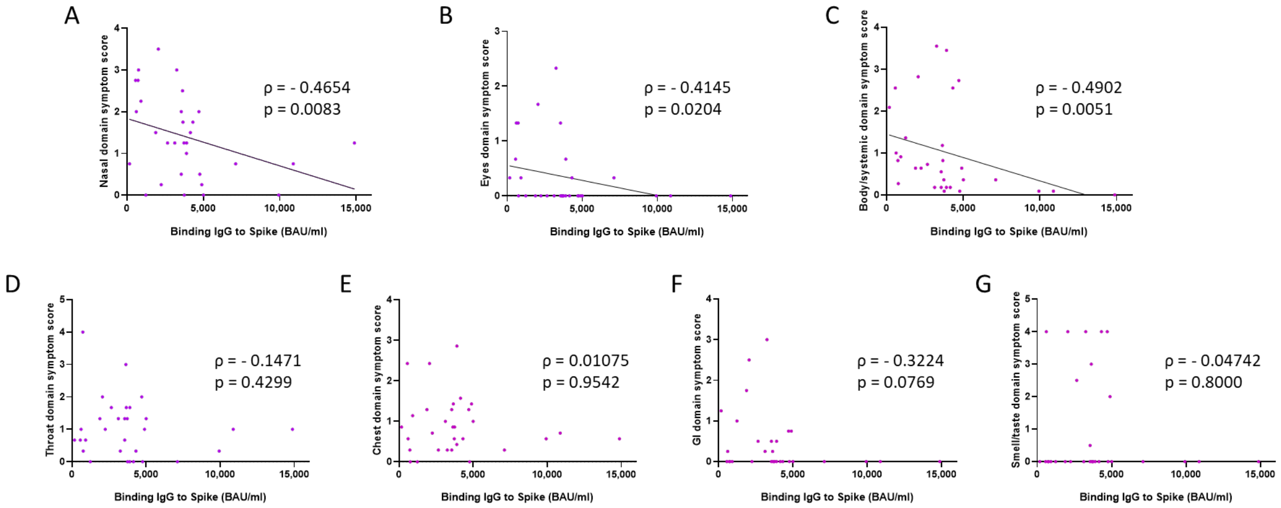


**Supplementary Figure 3. Correlation plots of anti-S (WT) IgG serum levels and domain symptom scores in the PVI group**

(A) Correlation between anti-S (WT) IgG serum levels (BAU/ml) and nasal domain symptom scores (Spearman $\rho = -0.4654$; $p = 0.0083$). (B) Correlation between anti-S (WT) IgG serum levels (BAU/ml) and eyes domain symptom scores (Spearman $\rho = -0.4145$; $p = 0.0204$). (C) Correlation between anti-S (WT) IgG serum levels (BAU/ml) and body/systemic domain symptom scores (Spearman $\rho = -0.4902$; $p = 0.0051$). (D) No correlation between anti-S (WT) IgG serum levels (BAU/ml) and throat domain symptom scores (Spearman $\rho = -0.1471$; $p = 0.4299$). (E) No correlation between anti-S (WT) IgG serum levels (BAU/ml) and chest domain symptom scores (Spearman $\rho = 0.01075$; $p = 0.9542$). (F) No correlation between anti-S (WT) IgG serum levels (BAU/ml) and gastrointestinal (GI) domain symptom scores (Spearman $\rho = -0.3224$; $p = 0.0769$). (G) No correlation between anti-S (WT) IgG serum levels (BAU/ml) and smell/taste domain symptom scores (Spearman $\rho = -0.04742$; $p = 0.8$). Dots indicate results from individual participants, n=32. * $p < 0.05$; ** $p < 0.01$; *** $p < 0.001$; **** $p < 0.0001$.

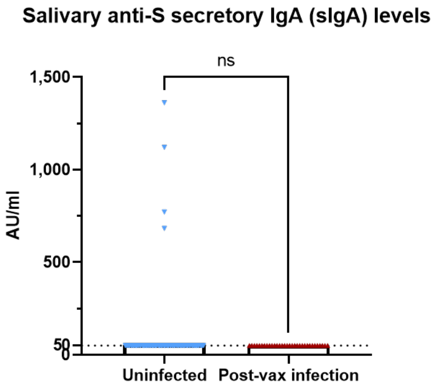


**Supplementary Figure 4. Saliva anti-S (WT) secretory IgA levels**

Saliva samples were obtained during the fall 2021 research clinic visit, at a median of 52.5 (IQR: 45.8 – 67.3) days after last immunization for the PVI group (n=32) and 46.5 (IQR: 32.8 – 58.3) days after last immunization for the uninfected group (n=143). A) Comparison of anti-S (WT) secretory IgA (sIgA) saliva levels between the uninfected group (n=143) and the PVI group (n=32). P values determined using the Mann-Whitney U test ($p = 0.5947$). Dots indicate results from individual participants and bars indicate geometric mean with 95% confidence intervals (CI). * $p < 0.05$; ** $p < 0.01$; *** $p < 0.001$; **** $p < 0.0001$.

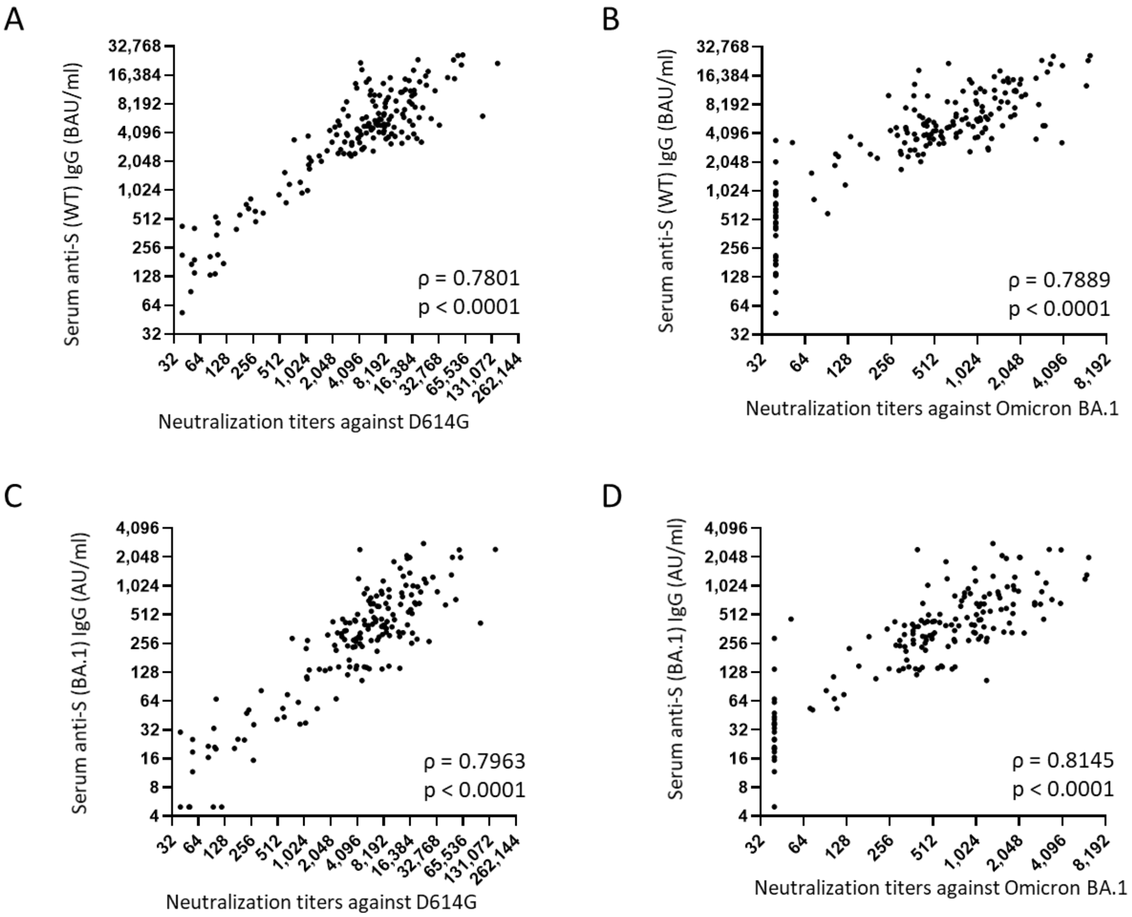


**Supplementary Figure 5. Correlation plots of serum binding antibody levels against WT and BA.1 Spike and pseudovirus neutralization antibody titers against WT and BA.1**

(A) Correlation between anti-S (WT) IgG serum levels (BAU/ml) and pseudovirus neutralization antibody titer ($ID_{50}$) against D614G (Spearman $\rho = 0.7801$; $p < 0.0001$). (B) Correlation between anti-S (WT) IgG serum levels (BAU/ml) and pseudovirus neutralization antibody titer ($ID_{50}$) against Omicron subvariant BA.1 (Spearman $\rho = 0.7889$; $p < 0.0001$). (C) Correlation between anti-S (BA.1) IgG serum levels (AU/ml) and pseudovirus neutralization antibody titer ($ID_{50}$) against D614G (Spearman $\rho = 0.7963$; $p < 0.0001$). (D) Correlation between anti-S (BA.1) IgG serum levels (AU/ml) and pseudovirus neutralization antibody titer ($ID_{50}$) against Omicron subvariant BA.1 (Spearman $\rho = 0.8145$; $p < 0.0001$). Dots indicate results from individual participants.

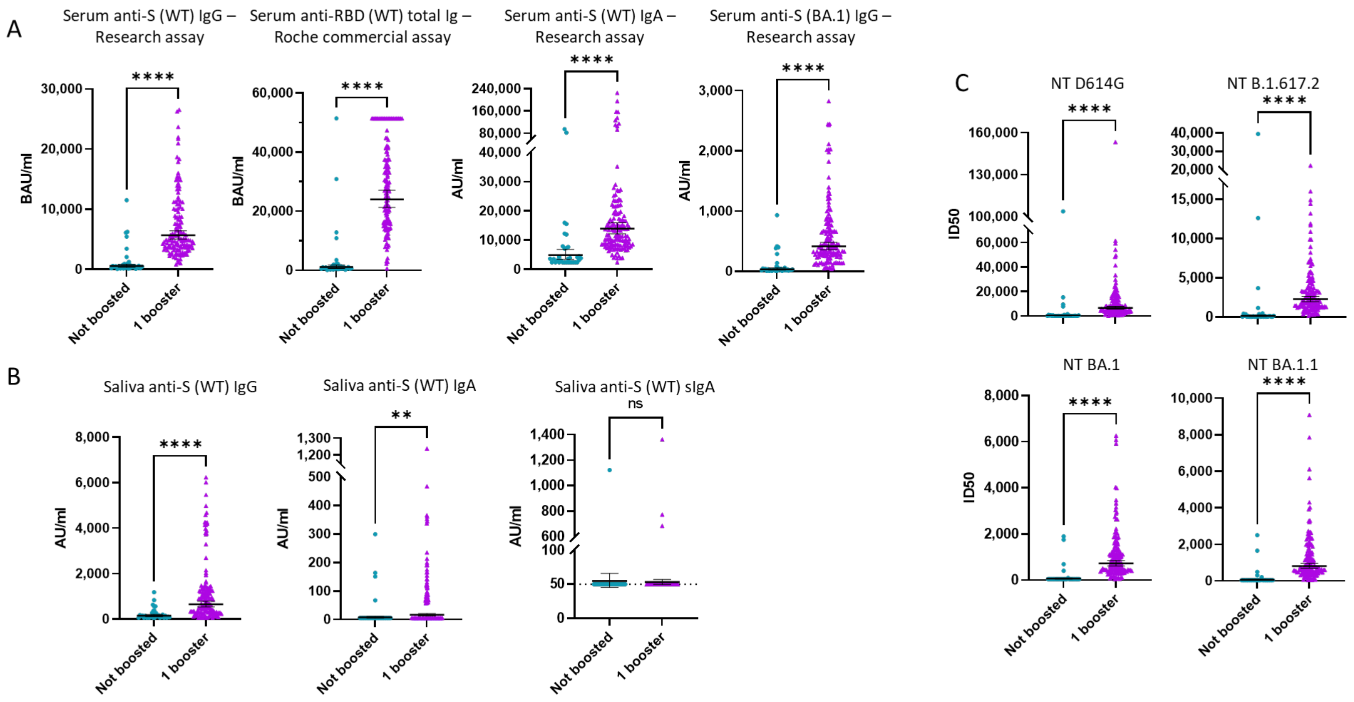


**Supplementary Figure 6. Comparison of binding antibody serum levels, binding antibody saliva levels, and pseudovirus neutralization antibody titers between unboosted and boosted participants**

(A) From left to right: comparison between unboosted and boosted participants of anti-S (WT) IgG serum levels (BAU/ml) measured with the research assay ($p < 0.0001$), anti-S (WT) total Ig serum levels (BAU/ml) measured with the Roche Elecsys® Anti-SARS-CoV-2 S assay ($p < 0.0001$), anti-S (WT) IgA serum levels (AU/ml) measured with the research assay ($p < 0.0001$), and anti-S (BA.1) IgG serum levels (AU/ml) measured with the research assay ($p < 0.0001$). (B) From left to right: comparison between unboosted and boosted participants of anti-S (WT) IgG saliva levels (AU/ml; $p < 0.0001$), anti-S (WT) IgA saliva levels (AU/ml; $p = 0.0079$), and anti-S (WT) sIgA saliva levels (AU/ml; $p = 0.7937$). (C) From left to right, top to bottom: comparison between unboosted and boosted participants of pseudovirus neutralization antibody titers ($ID_{50}$) against D614G ($p < 0.0001$), Delta variant B.1.617.2 ($p < 0.0001$), Omicron subvariant BA.1 ($p < 0.0001$), and Omicron subvariant BA.1.1 ($p < 0.0001$). P values determined using the Mann-Whitney U test. Dots indicate results from individual participants and bars indicate geometric mean with 95% CI. sIgA, secretory IgA. * $p < 0.05$; ** $p < 0.01$; *** $p < 0.001$; **** $p < 0.0001$.

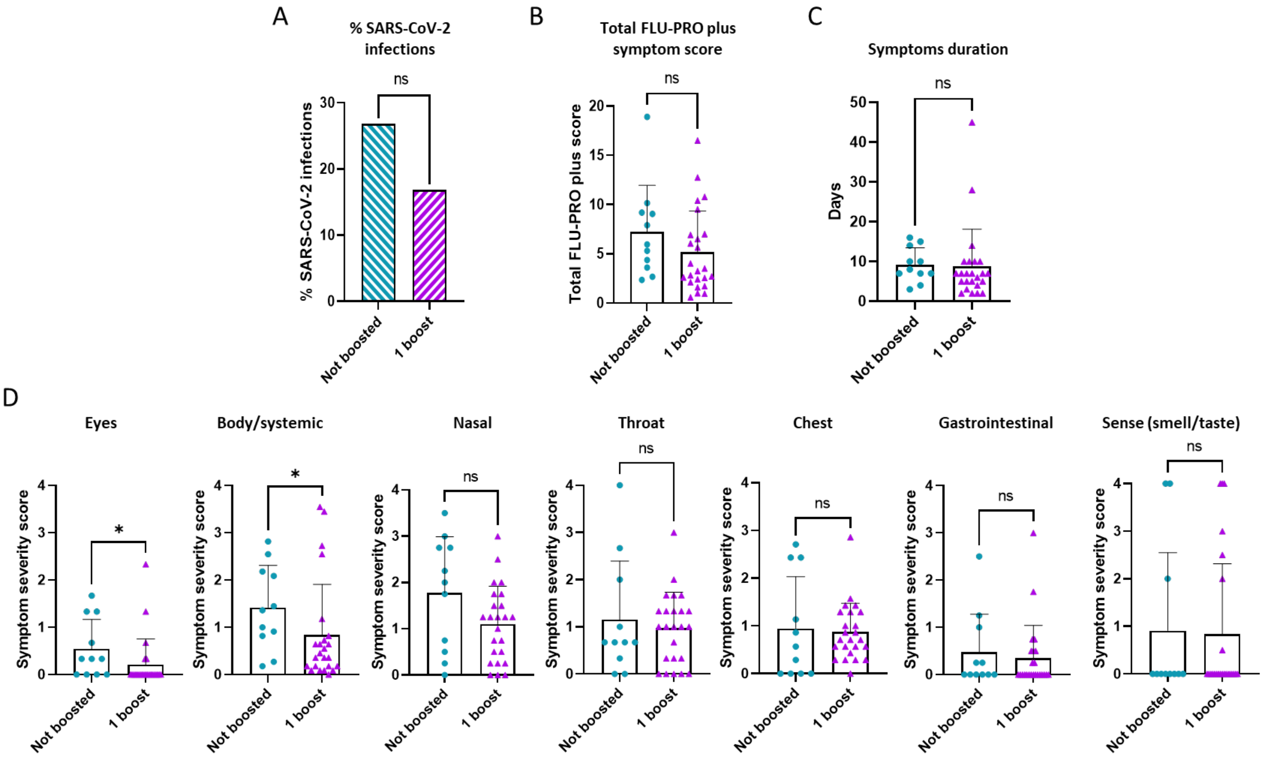


**Supplementary Figure 7. Impact of boosting status on SARS-CoV-2 infection's likelihood, overall symptoms severity and duration, as well as domain symptom severity**

(A) Percentage of infections in the unboosted versus boosted participants (p = 0.1776). (B) Total FLU-PRO plus symptom scores in the unboosted versus boosted participants (p = 0.1628). (C) Overall symptoms duration in the unboosted versus boosted participants (p = 0.1496). (D) Symptom severity scores for each symptom domain in the unboosted versus boosted participants (eyes, p = 0.016; body/systemic, p = 0.0237; nasal, p = 0.1072; throat, p = 0.8517; chest, p = 0.5088; gastrointestinal, p = 0.6377; sense, p > 0.9999). P values determined using the Mann-Whitney U test. Dots indicate results from individual participants and bars indicate mean with standard deviation. * p < 0.05; ** p < 0.01; *** p < 0.001; **** p < 0.0001.

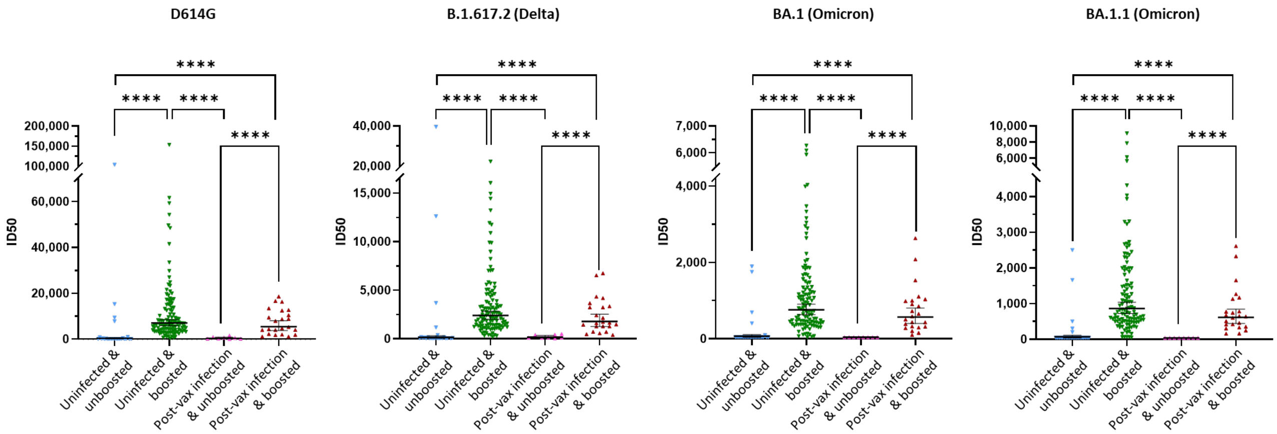


| GMT | | | | | | | | | | | | | | | |
|---|---|---|---|---|---|---|---|---|---|---|---|---|---|---|---|
| 252 | 7037 | 348 | 5432 | 144 | 2396 | 168 | 1788 | 69 | 757 | 40 | 567 | 70 | 865 | 40 | 611 |

**Supplementary Figure 8. Impact of boosting status on pseudovirus neutralization antibody titers in the uninfected versus post-vaccine infection group**

Pseudovirus neutralization ID50 titers against D614G, Delta variant B.1.617.2, and Omicron subvariants BA.1 and BA.1.1 for uninfected & unboosted participants (N=25), uninfected & boosted participants (N=115), PVI & unboosted participants (N=8), and PVI & boosted participants (N=23). P values determined using the Mann-Whitney U test, with a Bonferroni correction for 6 comparisons, alpha = 0.0083. Dots indicate results from individual participants and bars indicate geometric mean titers (GMT) with 95% CI, and GMTs are indicated. * $p < 0.05$; ** $p < 0.01$; *** $p < 0.001$; **** $p < 0.0001$.

| Work | | Home | |
|---|---|---|---|
| **1. Risk Exposure** | **2. Precautionary Measures** | **3. Risk Exposure** | **4. Precautionary Measures** |
| 1. Days working in the hospital (0-4) | 1. Consistent PPE use during patient interactions (0-4) | 1. Member of household tested COVID+ (0-4) | 1. Mask wearing outside of the home (grocery stores) (0-4) |
| 2. Direct contact with COVID+ patients (0-4) | 2. PPE use when not directly interacting with patients (0-4) | 2. Member of household had symptoms consistent with COVID and hasn't been tested (0-2) | 2. Mask wearing while outside doing solitary activities (0-4) |
| 3. Conducted high-risk activities on COVID+ patients (0-4) | 3. Consistency of applying hand sanitizer/washing hands before and after interacting with patients (0-4) | 3. Number of times in an out of house activity (grocery, restaurant, gym, bus…) (0-4 averaged for all 6 responses to this question) | 3. Consistently practicing social distancing outside home (0-4) |
| | | 4. Size and number of social gatherings (0-4 averaged for all 5 responses to this question) | 4. Disinfecting mail/packages delivered to home (0-4) |
| | | | 5. Type of mask typically used (1-4 averaged for all 4 responses to this question) |
| **Work Risk Score= (Total score/12)x100** | **Work Precautionary Score= (Total score/12)x100** | **Home Risk Score= (Total score/14)x100** | **Home Precautionary Score= (Total score/20)x100** |

**Supplementary Table 1. Calculation method for the Work Risk Score, Work Precautionary Score, Home Risk Score, and Home Precautionary Score**

These four scores are generated each month when a PASS participant fills out the "Risk Exposure, PPE Use, and Social Distancing" questionnaire. PPE, Personal Protective Equipment.

| Sample | Location | Collection date | # Raw Reads | # Joined Reads Mapped | Consensus genome length (nt) | Average Coverage (x) | Pangolin Lineage | Nextstrain Clade |
|---|---|---|---|---|---|---|---|---|
| 176 | Bethesda, MD | 01/2022 | 629,614 | 281,007 | 29,747 | 3,765 | BA.1.20 (Omicron) | 21K (Omicron)/high |
| 177 | Bethesda, MD | 02/2022 | 737,940 | 331,425 | 29,747 | 4,432 | BA.1.1 (Omicron) | 21K (Omicron)/high |
| 181 | Bethesda, MD | 12/2021 | 1,055,928 | 449,639 | 29,747 | 6,036 | BA.1.1 (Omicron) | 21K (Omicron)/high |
| 182 | Bethesda, MD | 01/2022 | 619,074 | 260,449 | 29,747 | 3,468 | BA.1.1 (Omicron) | 21K (Omicron)/high |
| 183 | Bethesda, MD | 01/2022 | 1,111,572 | 472,861 | 29,747 | 6,327 | BA.1.20 (Omicron) | 21K (Omicron)/high |
| 184 | Bethesda, MD | 12/2021 | 634,940 | 269,282 | 29,747 | 3,600 | BA.1.1 (Omicron) | 21K (Omicron)/high |
| 185 | Bethesda, MD | 12/2021 | 589,156 | 250,817 | 29,764 | 3,370 | AY.25 (Delta) | 21J (Delta)/high |
| 186 | Bethesda, MD | 12/2021 | 1,001,634 | 412,655 | 29,708 | 5,563 | BA.1.18 (Omicron) | 21K (Omicron)/high |
| 187 | Bethesda, MD | 12/2021 | 643,086 | 271,882 | 29,747 | 3,622 | BA.1.15 (Omicron) | 21K (Omicron)/high |
| 188 | Bethesda, MD | 12/2021 | 714,018 | 299,757 | 29,747 | 3,960 | BA.1.1 (Omicron) | 21K (Omicron)/high |
| 189 | Bethesda, MD | 12/2021 | 717,104 | 307,496 | 29,708 | 4,105 | BA.1.18 (Omicron) | 21K (Omicron)/high |
| 190 | Bethesda, MD | 12/2021 | 465,830 | 197,371 | 29,747 | 2,587 | BA.1.1 (Omicron) | 21K (Omicron)/high |
| 191 | Bethesda, MD | 01/2022 | 769,504 | 326,309 | 29,747 | 4,357 | BA.1.19 (Omicron) | 21K (Omicron)/high |

**Supplementary Table 2. Sequencing statistics for 13 SARS-CoV-2 post-vaccine infections**

All samples produced coding complete genomes. Viral Amplicon Illumina Workflow 2.3 was used to collate and analyze SARS-CoV-2 genomes from the resulting sequencing reads. Consensus genomes were generated when possible. Lineage determination of consensus genomes was conducted using Pangolin (Phylogenetic Assignment of Named Global Outbreak LINeages; v4.1.2). Nextstrain clades were determined by Nextclade CLI 2.4.0, Nextalign CLI 1.10.1. Nextstrain overall sequence QC scores of ‘bad’, ‘mediocre’, and ‘good’ were translated into ‘low’, ‘medium’, and ‘high’ for confidence of clade assignment.

| | Unadjusted | |
|---|---|---|
| | OR (95% CI) | P-value |
| Age | 0.973 (0.826-1.147) | 0.746 |
| Female | 0.406 (0.185-0.893) | 0.025 |
| Serum anti-S (WT) IgG (BAU/ml) | 0.703 (0.527-0.937) | 0.016 |
| Serum anti-S (BA.1) IgG (AU/ml) | 0.811 (0.616-1.067) | 0.134 |
| Serum anti-S (WT) IgA (AU/ml) | 0.489 (0.180-1.329) | 0.161 |
| Roche Serum anti-RBD (WT) total Ig (BAU/ml) | 0.479 (0.177-1.299) | 0.148 |
| Neutralizing titer D614G (ID50) | 0.480 (0.197-1.166) | 0.105 |
| Neutralizing titer B.1.617.2 (ID50) | 0.498 (0.236-1.050) | 0.067 |
| Neutralizing titer BA.1 (ID50) | 0.694 (0.477-1.010) | 0.056 |
| Neutralizing titer BA.1.1 (ID50) | 0.573 (0.341-0.961) | 0.035 |
| Saliva anti-S (WT) IgG (AU/ml) | 1.051 (0.884-1.249) | 0.576 |
| Saliva anti-S (WT) IgA (AU/ml) | 0.867 (0.521-1.443) | 0.583 |
| Work risk score (%) | 1.074 (0.921-1.253) | 0.362 |
| Work precautionary score (%) | 0.998 (0.844-1.180) | 0.979 |
| Home risk score (%) | 2.144 (1.645-2.794) | 0.000 |
| Home risk score 1-2 (%) | 1.760 (1.472-2.103) | 0.000 |
| Home risk score 3-4 (%) | 1.044 (0.869-1.253) | 0.647 |
| Home precautionary score (%) | 1.009 (0.842-1.209) | 0.923 |
| | **Adjusted for age, sex and home risk score** | |
| | **OR (95% CI)** | **P-value** |
| Serum anti-S (WT) IgG (BAU/ml) | 0.673 (0.477-0.950) | 0.025 |
| Serum anti-S (BA.1) IgG (AU/ml) | 0.785 (0.572-1.079) | 0.136 |
| Serum anti-S (WT) IgA (AU/ml) | 0.428 (0.109-1.680) | 0.224 |
| Roche Serum anti-RBD (WT) total Ig (BAU/ml) | 0.426 (0.109-1.662) | 0.219 |
| Neutralizing titer D614G (ID50) | 0.350 (0.109-1.124) | 0.078 |
| Neutralizing titer B.1.617.2 (ID50) | 0.409 (0.151-1.111) | 0.080 |
| Neutralizing titer BA.1 (ID50) | 0.705 (0.445-1.117) | 0.137 |
| Neutralizing titer BA.1.1 (ID50) | 0.511 (0.256-1.018) | 0.056 |
| Saliva anti-S (WT) IgG (AU/ml) | 1.014 (0.800-1.285) | 0.908 |
| Saliva anti-S (WT) IgA (AU/ml) | 0.851 (0.414-1.749) | 0.661 |

**Supplementary Table 3. Numeric values of point estimates and 95% CI for the odds ratios unadjusted and adjusted for non-immunological factors**
